# Processing Speed Accounts for the Discriminative Power of a Smartphone Go/No-Go Task in Multiple Sclerosis

**DOI:** 10.64898/2026.09.09.26362640

**Authors:** Peter Kosa, Amir Moghadam Ahmadi, Marie Kanu, Yolanda Mejia, Emmanuel Mekasha, Caledonia Steltzner, Bibiana Bielekova

**Affiliations:** Neuroimmunological Diseases Section, Laboratory of Clinical Immunology and Microbiology, National Institute of Allergy and Infectious Diseases, National Institutes of Health, Bethesda, MD, USA

**Keywords:** cognitive decline, digital biomarkers, phone app, multiple sclerosis, Go/No-Go test

## Abstract

**Background:** Identifying and longitudinally measuring cognitive decline remains challenging because traditional assessments rely largely on episodic, in-person administration of analog scales by trained examiners. Smartphone technology provides a scalable alternative capable of extracting distinct digital biomarkers from continuous sensor and event streams. Following the integration of several cognitive modules into the self-administered Neurological Functions Test Suite (NeuFun-TS), this study evaluates the clinical utility and incremental validity of a final candidate module: an equiprobable (50/50) continuous vigilance Go/No-Go paradigm.

**Methods:** We analyzed 3,270 quality-controlled trials from 303 participants, focusing primary analyses on 488 first-trial observations across 34 healthy donors (HD) and 213 multiple sclerosis (MS) participants. From the raw event stream, we screened 16 candidate metrics and retained 10 based on their intrinsic psychometric properties. Using these 10 metrics, we evaluated diagnostic group separation, test-retest reliability, correlations with clinical and magnetic resonance imaging (MRI) benchmarks, and incremental validity over existing digital modules: the randomized Symbol Digit Modalities Test (rSDMT) and the Motor Sequencing Test (MST), using paired observations.

**Results:** Although nine of the 10 outcomes significantly separated HD from MS and seven separated relapsing-remitting from progressive MS subtypes (all adjusted p < 0.05), discriminative power was driven almost entirely by response latency rather than commission errors or inhibitory metrics. Multi-predictor composite scores offered no diagnostic advantage over reaction time alone. Furthermore, the Go/No-Go task demonstrated poor-to-moderate longitudinal test-retest reliability and failed to provide incremental clinical value over existing modules.

**Conclusion:** Although the smartphone-based Go/No-Go task reflects processing speed impairment in MS, its discriminative capability is redundant with simpler reaction time metrics already implemented in the platform. Consequently, the Go/No-Go module was excluded from NeuFun-TS to minimize patient testing burden without compromising diagnostic sensitivity.

## 1. Introduction

Accurately monitoring disease progression in chronic neurological diseases such as multiple sclerosis (MS) requires frequent, objective, and comprehensive assessment of neurological functions. Unlike platforms designed to supplement episodic clinic visits or import isolated analog tests onto a screen^1–4^, the Neurological Functions Test Suite (NeuFun-TS^5–11^) aims to recreate all aspects of the formal neurological examination that can be measured via smartphone. NeuFun-TS^5–12^ differentiates itself on five foundational principles: 1. Systematic mapping to discrete, anatomically defined neurological subsystems (visual, pyramidal, cerebellar, sensory, and cognitive); 2. Rigorous multimodal validation against anatomically/functionally corresponding results from concurrent clinician-derived disability scales (i.e., relevant subdomains of the d0igitalized Neurological Examination [NeurEx^TM^]^13^, Expanded Disability Status Scale [EDSS]^14^ and Combinatorial, Weight-Adjusted Disability Scale [CombiWISE]^15^) and MRI metrics of brain and spinal cord tissue destruction (i.e., Composite MRI Scale of CNS tissue destruction [COMRIS-CTD]^16^); 3. Mandatory, unbiased validation where predictive models are frozen on training data and tested on independent, held-out cohorts; 4. Isolation of physiological confounders, such as separating disease-specific dysfunction from natural aging^11^ and decomposing cognitive processing speed from motor disability^7^; and 5. Unsupervised closed-loop quality assurance alongside iterative suite optimization, wherein successful testing paradigms may be further enhanced^8,11^, while tests that prove unreliable or redundant (such as spiral tracing^12^, pronator drift^11^, and verbal memory^9^) are systematically retired.

Cognitive impairment, predominantly slowing of information processing speed (IPS)^17^, attentional deficits, and executive dysfunction^18,19^, is a major contributor to disability in MS^20^. To capture real-world cognitive-motor speed and sustained attention, we implemented an equiprobable (50/50) Go/No-Go choice-reaction task that we’ll abbreviate to 50/50GNG. Unlike traditional skewed paradigms (e.g., 80% Go/20% No-Go) that evaluate response inhibition (a process localized to the right-inferior frontal gyrus and pre-supplementary motor area connected to the subthalamic nucleus via a fronto-basal ganglia network^21^), an equiprobable design shifts cognitive demand toward sustained attention and choice reaction time under time pressure, which we hypothesized would be more sensitive to MS-related impairment.

Because NeuFun-TS aims to maximize information per unit of patient burden, any candidate test must demonstrate incremental clinical value over existing modules. In this study, we evaluated the clinical utility, psychometric properties, and redundancy of the 50/50GNG against established NeuFun-TS cognitive and motor benchmarks, specifically the randomized Symbol Digit Modalities Test (rSDMT^7^), the Motor Sequencing Test (MST^10^) and the Spatial Memory Test^9^.

## 2. Methods

### 2.1 Participants

Participants were enrolled into two National Institutes of Health (NIH) protocols, NCT00794352 (“Comprehensive Multimodal Analysis of Neuroimmunological Diseases of the CNS”) and NCT03109288 (“Targeting Residual Activity by Precision, Biomarker-Guided Combination Therapies of MS”), both approved by the NIH Institutional Review Board. All participants provided written or electronic informed consent. Smartphone testing occurred during routine study visits between February 2021 and June 2026.

We prospectively streamed raw data to a secure database under assigned alphanumeric codes and downloaded them in bulk for the analysis. After quality control (QC, see below), we unblinded diagnoses from the research database as healthy donor (HD), relapsing-remitting MS (RR-MS), secondary progressive MS (SP-MS), or primary progressive MS (PP-MS). For ordinal analyses, we combined the two progressive subtypes (P-MS). We held out a separate cohort of participants with non-MS neurological diagnoses, including non-inflammatory neurological disease (NIND), other inflammatory neurological disease (OIND), and clinically/radiologically isolated syndrome (CIS/RIS; subjects who did not yet convert to definite MS), from all primary analyses, reserving them for the discriminant validity assessment reported in Supplementary Material.

Figure 1 shows the demographic and clinical characteristics of all seven diagnostic groups - age, sex, disease duration, EDSS score, and the trials and first-trial observations each group contributes. Supplementary Table S1 gives the same data numerically.

**Figure 1.**
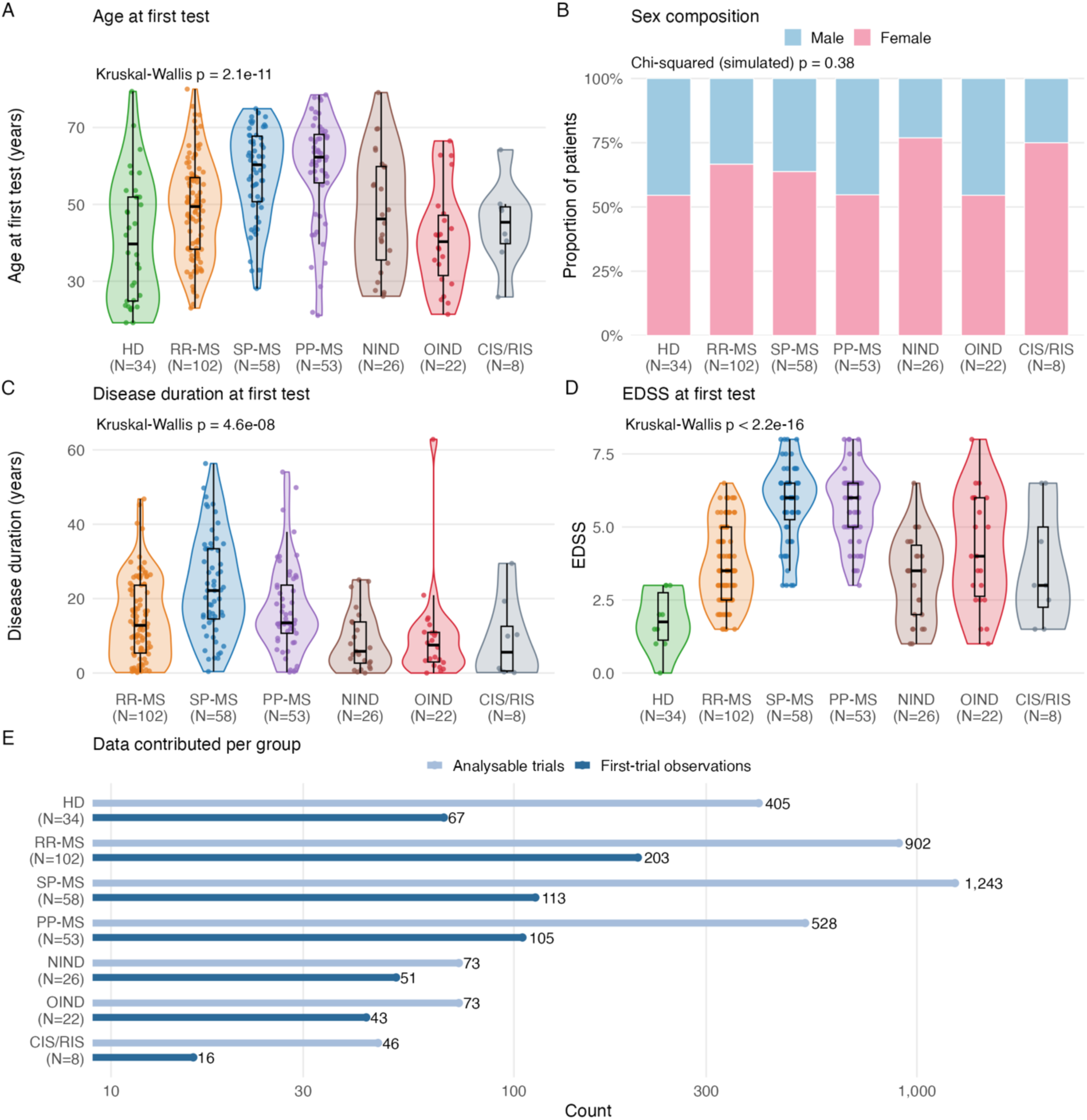
Cohort composition across the seven diagnostic groups: the four groups of the primary analysis (HD, RR-MS, SP-MS, PP-MS) and the three non-MS groups held out of it (NIND, OIND, CIS/RIS). Supplementary Table S1 gives the same data numerically. **(A)** Age at first analyzable trial. **(B)** Sex composition as a stacked proportion, male in blue and female in pink. **(C)** Disease duration at first analyzable trial; HD have no onset date and are omitted. **(D)** EDSS from the closest clinician exam within ±7 days of that trial. **(E)** Analyzable trials and first-trial observations per group, on a logarithmic axis. In A, C and D violins are trimmed to each group’s observed range, boxes give the median and quartiles, and every participant is plotted as a point; groups with fewer than three values are drawn as points without a violin. Each axis label carries the group’s N in participants. Tests are descriptive and unadjusted: Kruskal-Wallis across the seven groups in A, C and D, and a chi-squared test with a simulated p in B. See Section 3.1. Abbreviations: CIS/RIS, clinically or radiologically isolated syndrome; EDSS, Expanded Disability Status Scale; HD, healthy donor; MS, multiple sclerosis; N, number of participants; NIND, non-inflammatory neurological disease; OIND, other inflammatory neurological disease; PP-MS, primary progressive multiple sclerosis; RR-MS, relapsing-remitting multiple sclerosis; SP-MS, secondary progressive multiple sclerosis.

### 2.2 Sustained attention and choice-reaction task under pressure

We administered the 50/50GNG choice-reaction task on Google Pixel XL smartphones as a module within NeuFun-TS. Participants performed the task once with each hand. A single blue circle (“Go”) required a two-finger tap, whereas a pair of blue circles (“No-Go”) required withholding the response (Figure 2A–C). The application displayed each stimulus for up to 900 milliseconds (ms), followed by a 100 ms blank interval. The screen dismissed a tapped stimulus upon finger release, allowing faster participants to complete more stimulus cycles within the fixed trial time of approximately 30 seconds (s).

**Figure 2.**
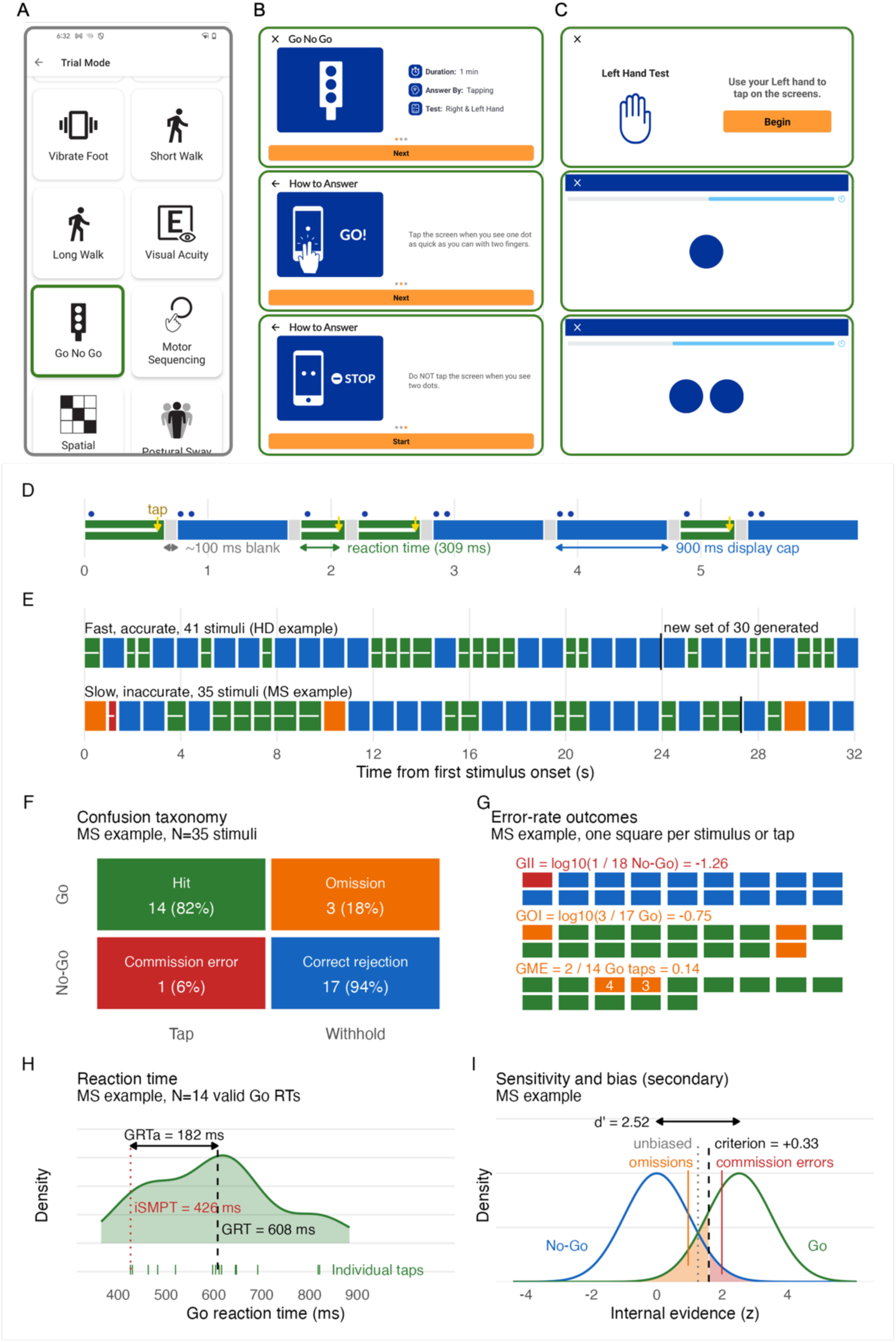
The Go/No-Go task as administered and as recorded. **(A–C)** Screen captures from the study build in the order the participant meets them: the test suite with the Go/No-Go module outlined, the instruction screens defining the stimulus-response rules, and the hand prompt with the Go and No-Go stimuli as they appear during the timed trial. **(D–E)** Two complete recorded trials on a common wall-clock axis, with panel D expanding the first 5.4 s of the fast trial. Dots above each rectangle are the stimulus as presented; the rectangle’s color is the response the trial was scored as; white bars mark reaction times and gray bars the ∼100 ms blank interval. The fast performer (HD, 45 y) receives 41 stimuli in 32.2 s with no errors; the slow performer (SP-MS, 68 y) receives 35 stimuli in 32.0 s, misses 3 of 17 Go stimuli and commits one commission error. Both lanes span the same wall-clock time. **(F–I)** The five primary outcomes and the two signal-detection parameters, all defined on the slow trial of panel **E**: the stimulus-by-response taxonomy **(F)**, the three error-rate outcomes **(G)**, GRT, iSMPT and their difference GRTa over a kernel density of this trial’s valid Go reaction times **(H)**, and d′ and criterion placed to reproduce this trial’s hit and commission-error rates exactly **(I)**. Abbreviations: d′, sensitivity; GII, Go/No-Go inhibition impairment; GME, Go/No-Go method error rate; GOI, Go/No-Go omission impairment; GRT, Go reaction time; GRTa, Go/No-Go adjusted reaction time; HD, healthy donor; iSMPT, individualized sensory-motor processing threshold; MS, multiple sclerosis; RT, reaction time; SP-MS, secondary progressive multiple sclerosis.

The task drew stimuli without replacement from a balanced set of 30 items (15 Go, 15 No-Go), reshuffling and repeating the deck until the time budget expired. This equiprobable design ensures the task measures continuous attention, rather than the inhibition of a habitual response tested by a skewed stimulus ratio. We empirically verified the composition of the stimulus stream from the recorded events, confirming the balanced design and establishing that the total stimulus count per trial functions as an inverse index of processing speed (Supplementary Methods SM1).

### 2.3 Quality control (QC) and analysis dataset

We applied four technical-validity criteria for trial-level QC: completeness (≥ 30 stimuli), duration (within 28–40 s), pacing (mean wall-clock time per stimulus ≥ 500 ms), and response validity (≤ 50% of taps below the 100 ms floor; see Supplementary Methods SM2). These criteria removed 140 of 3,444 extracted trials (4.1%).

We then applied a subject-level gate for task engagement, excluding trials where responses carried no information about the stimulus (Supplementary Methods SM2). This step removed an additional 34 of 3,304 trials (1.0%). No participant lost all data to these criteria, and we confirmed that our filtering did not select against participants with slower processing speeds.

For all cross-sectional comparisons, our primary analysis dataset consisted of the first valid trial per participant per hand to ensure statistical independence, yielding 488 observations across the primary MS and HD cohorts. For repeated-measures analyses, we used all 3,270 valid trials modeled with a participant-level random intercept.

### 2.4 Outcome derivation

We computed every outcome within a single trial. We defined reaction time as the latency from stimulus onset to the first screen interaction (Figure 2D-E). We counted a Go reaction time as valid if the participant tapped the Go stimulus and the latency fell between 100 and 2,000 ms; we excluded faster responses as anticipations or touch artifacts and slower responses as inattention rather than motor performance (see Supplementary Methods SM3).

We classified responses into four categories (Figure 2F): hits (tapped Go stimuli), omissions (untapped Go stimuli), commission errors (tapped No-Go stimuli at or above the 100 ms floor), and correct rejections (untapped No-Go stimuli). We calculated the hit rate as hits divided by presented Go stimuli, the commission-error rate as commission errors divided by presented No-Go stimuli, and the omission rate as one minus the hit rate.

We computed 16 candidate outcomes. Before assessing associations with any diagnosis, we screened candidate outcomes based on their intrinsic measurement properties (i.e., estimability, coverage, reliability, and redundancy). This process retained 10 outcomes (five primary, five secondary) that served as the denominator for multiple-testing corrections. Table 1 provides details for all 16 outcomes and Supplementary Methods SM3 gives details of their calculations.

**Table 1.**
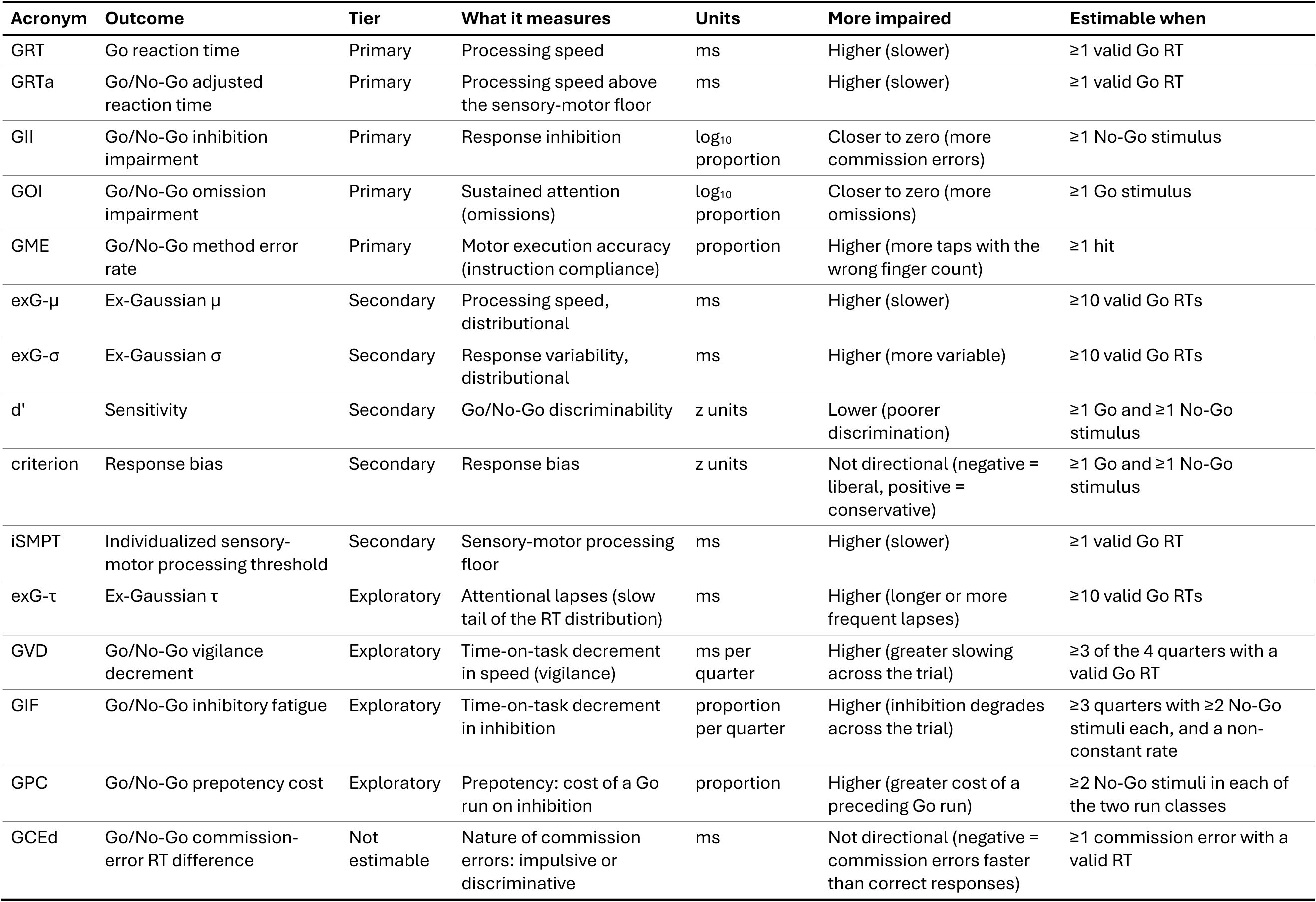
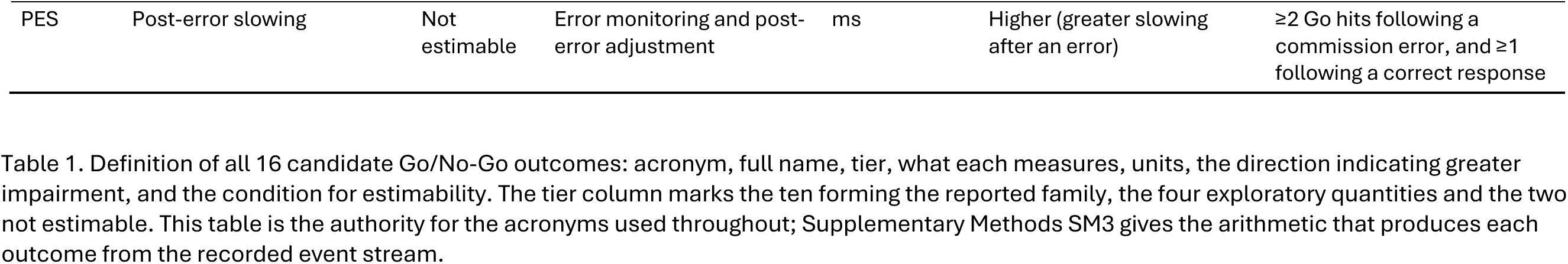
Definition of all 16 candidate Go/No-Go outcomes: acronym, full name, tier, what each measures, units, the direction indicating greater impairment, and the condition for estimability. The tier column marks the ten forming the reported family, the four exploratory quantities and the two not estimable. This table is the authority for the acronyms used throughout; Supplementary Methods SM3 gives the arithmetic that produces each outcome from the recorded event stream.

| Acronym | Outcome | Tier | What it measures | Units | More impaired | Estimable when |
| --- | --- | --- | --- | --- | --- | --- |
| GRT | Go reaction time | Primary | Processing speed | ms | Higher (slower) | ≥1 valid Go RT |
| GRTa | Go/No-Go adjusted reaction time | Primary | Processing speed above the sensory-motor floor | ms | Higher (slower) | ≥1 valid Go RT |
| GII | Go/No-Go inhibition impairment | Primary | Response inhibition | log<sub>10</sub> proportion | Closer to zero (more commission errors) | ≥1 No-Go stimulus |
| GOI | Go/No-Go omission impairment | Primary | Sustained attention (omissions) | log<sub>10</sub> proportion | Closer to zero (more omissions) | ≥1 Go stimulus |
| GME | Go/No-Go method error rate | Primary | Motor execution accuracy (instruction compliance) | proportion | Higher (more taps with the wrong finger count) | ≥1 hit |
| exG-μ | Ex-Gaussian μ | Secondary | Processing speed, distributional | ms | Higher (slower) | ≥10 valid Go RTs |
| exG-σ | Ex-Gaussian σ | Secondary | Response variability, distributional | ms | Higher (more variable) | ≥10 valid Go RTs |
| d' | Sensitivity | Secondary | Go/No-Go discriminability | z units | Lower (poorer discrimination) | ≥1 Go and ≥1 No-Go stimulus |
| criterion | Response bias | Secondary | Response bias | z units | Not directional (negative = liberal, positive = conservative) | ≥1 Go and ≥1 No-Go stimulus |
| iSMPT | Individualized sensory-motor processing threshold | Secondary | Sensory-motor processing floor | ms | Higher (slower) | ≥1 valid Go RT |
| exG-τ | Ex-Gaussian τ | Exploratory | Attentional lapses (slow tail of the RT distribution) | ms | Higher (longer or more frequent lapses) | ≥10 valid Go RTs |
| GVD | Go/No-Go vigilance decrement | Exploratory | Time-on-task decrement in speed (vigilance) | ms per quarter | Higher (greater slowing across the trial) | ≥3 of the 4 quarters with a valid Go RT |
| GIF | Go/No-Go inhibitory fatigue | Exploratory | Time-on-task decrement in inhibition | proportion per quarter | Higher (inhibition degrades across the trial) | ≥3 quarters with ≥2 No-Go stimuli each, and a non-constant rate |
| GPC | Go/No-Go prepotency cost | Exploratory | Prepotency: cost of a Go run on inhibition | proportion | Higher (greater cost of a preceding Go run) | ≥2 No-Go stimuli in each of the two run classes |
| GCEd | Go/No-Go commission-error RT difference | Not estimable | Nature of commission errors: impulsive or discriminative | ms | Not directional (negative = commission errors faster than correct responses) | ≥1 commission error with a valid RT |
| PES | Post-error slowing | Not estimable | Error monitoring and post-error adjustment | ms | Higher (greater slowing after an error) | $\geq 2$ Go hits following a commission error, and $\geq 1$ following a correct response |

The five primary outcomes are: the Go reaction time (GRT) to measure IPS; the Go adjusted reaction time (GRTa) to measure IPS above the sensory-motor floor; the Go/No-Go inhibition impairment (GII) to quantify response inhibition; the Go/No-Go omission impairment (GOI) to capture sustained attention through omissions; and the Go/No-Go method error rate (GME) to assess motor execution accuracy and instruction compliance. The five secondary outcomes are: the ex-Gaussian μ (exG-μ) and ex-Gaussian σ (exG-σ) to describe processing speed and response variability distributionally; two signal-detection parameters, sensitivity (d′) and response bias (criterion), to separate 50/50GNG discriminability from response tendency; and the individualized sensory-motor processing threshold (iSMPT) to establish the sensory-motor processing floor for each subject.

### 2.5 Correlations with clinical outcomes and assessing redundancy against digital comparators

To conduct the redundancy analysis, we evaluated the 50/50GNG against validated modules already deployed in NeuFun-TS. Our primary functional benchmarks were the digital MST^10^, using its validated two-predictor composite score (MSC-2), the Spatial memory composite^9^ and the rSDMT^7^.

The clinical comparators were four global disability scores: EDSS^14^, Scripps Neurological Rating Scale (SNRS^22^), CombiWISE^15^, and the total of the continuous disability scale computed by the NeurEx App^13^ (NeurEx total), together with four NeurEx subscores: upper extremity strength, pyramidal function, coordination and the cognitive panel^13^. The imaging comparators came from the COMRIS^16^ model: its global CNS tissue-destruction score (COMRIS-CTD) and nine regional metrics. With the rSDMT, the MSC-2 and the spatial memory composite, this gives 21 comparator measures in three blocks: eight clinical, ten imaging and three digital. The bidirectional variance analysis of Section 3.6 used four outcomes (one per measurement modality): CombiWISE (the examining neurologist), the rSDMT (the smartphone test of IPS), COMRIS-CTD (the scanner) and the 9-hole peg test (9HPT, a stopwatch; scored as the log of the average of 2 completion times, matched to the hand tested. Failure to complete the test was assigned sentinel values of 777 s). The 9HPT enters that analysis only and is not one of the selected 21 correlation comparators.

We matched every comparator temporally to the task trial date within ±7 days and used the same window for the paired dataset of the redundancy analysis. Nearly all matches are the same day. We restricted all correlation analyses to MS, leaving N=378 clinical observations from 199 patients, N=364 imaging observations from 191 patients, and N=241 digital observations from 129 patients.

### 2.6 Statistical analyses

We designed our statistical plan first to validate the task’s psychometric properties and then to assess its incremental diagnostic value for inclusion in NeuFun-TS.

We assessed group differences using Kruskal-Wallis tests with Benjamini-Hochberg false-discovery rate (FDR) correction across the ten reported outcomes. To evaluate disease staging (HD < RR-MS < P-MS), we constructed a multivariable 50/50GNG composite (GNG-C) using ordinal logistic regression on a 70/30 training and validation split of the MS cohort (Supplementary Methods SM4). We determined effect sizes for group contrasts as rank-biserial correlations (r) derived from the Mann-Whitney U statistic, signed so that positive values indicate the more impaired group scores higher on that outcome’s own scale.

We evaluated reliability across two distinct timescales: within-session internal consistency using the Spearman-Brown corrected correlation (r_SB_) between odd- and even-numbered stimuli, and between-session test-retest reliability using the two-way consistency, single-measures intraclass correlation coefficient (ICC), across the first three trials per patient-hand within a 90-day window (Supplementary Methods SM5).

To assess test redundancy against other existing tests, we performed three pre-specified analyses: 1. Incremental staging: We compared the held-out concordance index (median over 500 patient-level splits) of three nested ordinal models: MST alone, 50/50GNG alone, and their combination, refitting each model within every training split. 2. Partial correlation: We re-evaluated the correlations between 50/50GNG metrics and clinical outcomes after adjusting for MST speed measures, determining the proportion of the raw association that remained. 3. Unique variance in both directions: We used nested linear models to quantify the incremental explained variance (ΔR²) that each 50/50GNG outcome added over the MST baseline for the four outcomes named in Section 2.5 (Supplementary Methods SM6).We then reversed this comparison on identical rows to measure the variance MST added over 50/50GNG. Evaluating this bidirectional asymmetry distinguishes true independent signal from the partial absorption of correlated metrics.

Finally, we assessed discriminant validity by applying GNG-C, with the coefficients of the staging model held fixed, to the held-out non-MS groups, comparing all seven diagnostic groups by Kruskal-Wallis test and all 21 group pairs by Wilcoxon rank-sum test with Benjamini-Hochberg correction across the full set of 21 (Supplementary Methods SM7).

We conducted all analyses in R version 4.5.1^23^. The analysis code and derived datasets required to reproduce all results, figures and tables will be openly available at https://github.com/Bielekova-Lab/neufun-go-no-go upon publication.

## 3. Results

### 3.1 Cohort and task implementation

Across all cohorts, 303 participants completed at least one analyzable trial (Figure 1; Supplementary Table S1). We limited primary analyses to the 247 HD and MS participants, contributing 488 first-trial patient-hand observations across the four main diagnostic groups: HD (N=67 hands), RR-MS (N=203), SP-MS (N=113), and PP-MS (N=105). We held out an additional 110 observations from three non-MS neurological disease groups (NIND, OIND, CIS/RIS) for discriminant validity testing (Supplementary Figure S1, Supplementary Results SR1). The rigorous QC pipeline excluded 4.1% of all extracted trials for technical invalidity and an additional 1.0% for non-discriminating responses, with no evidence of bias against participants with severe impairment (Supplementary Results SR2).

Our empirical analysis of the recorded event streams confirmed that the task operates as a continuous attention paradigm. Across all 3,270 analyzable trials, the stimulus deck was perfectly balanced (15 Go and 15 No-Go items in the first 30 stimuli) with an unpredictable, random-shuffle presentation order. This equiprobable (50/50) design requires continuous cognitive engagement and active decision-making rather than the suppression of an incorrect, habitual response (i.e., response inhibition). Consequently, the total number of stimuli a participant completed within the fixed 30-second trial served as a reliable inverse index of their processing speed (ρ = -0.87 with GRT; Supplementary Results SR3).

### 3.2 The 50/50GNG is sensitive to disease-related cognitive impairment, but the IPS dominates its performance

We found that the task successfully detected disease-related impairment: across first-trial data (N = 488), nine of the ten reported outcomes differed significantly across the four diagnostic groups (Kruskal-Wallis, Benjamini-Hochberg-adjusted p<0.05), with GME being the sole exception (Figure 3A; Supplementary Table S2). Resolving each omnibus test into its six pairwise comparisons (Mann-Whitney, Holm-adjusted within an outcome and interpreted only where the omnibus test survived multiple testing correction), nine of the ten outcomes separated HD from at least one MS subtype and seven separated RR-MS from at least one progressive subtype, while none separated SP-MS from PP-MS (Supplementary Table S3).

**Figure 3.**
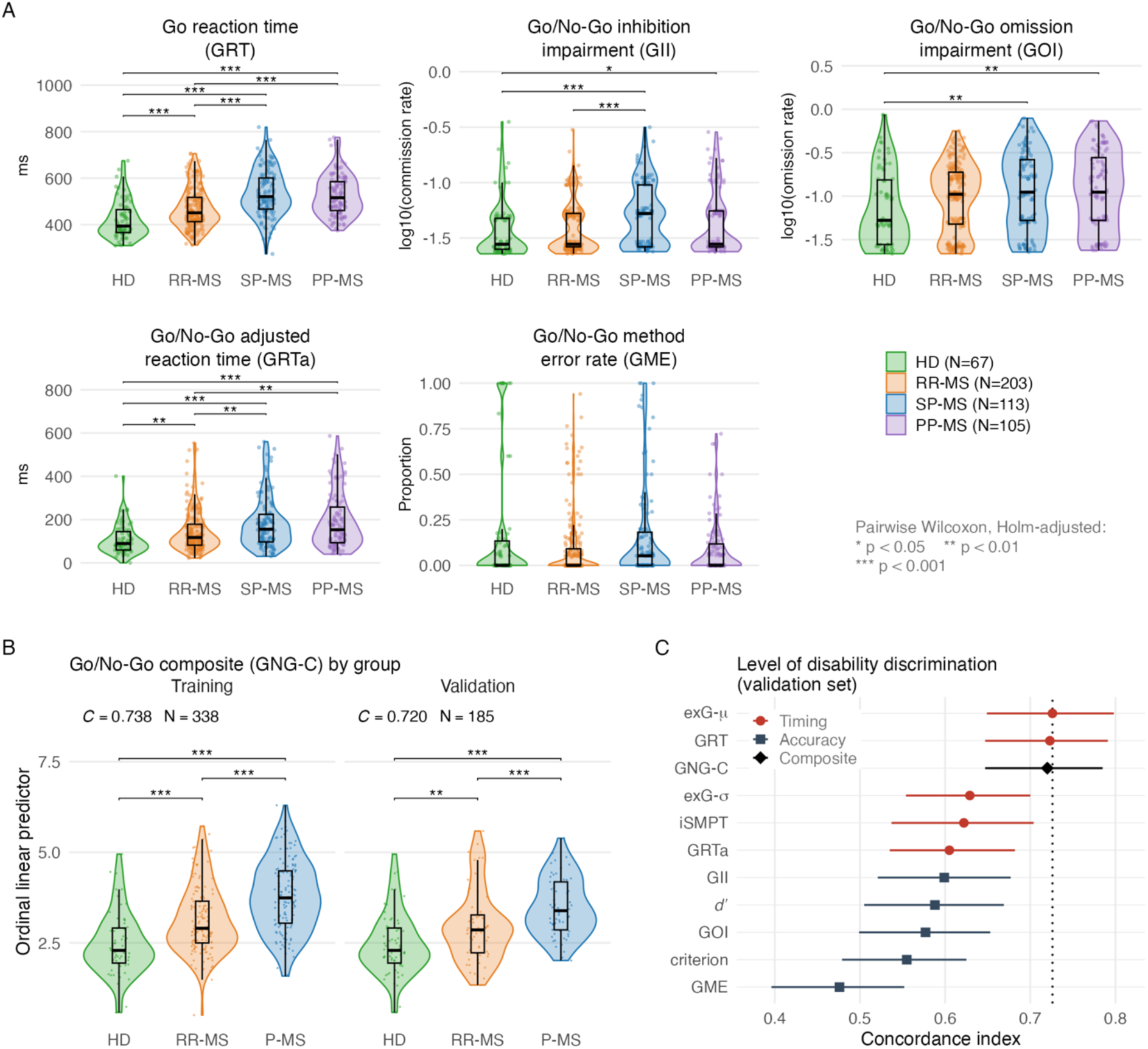
Group separation is carried by processing speed. **(A)** The five primary outcomes by diagnostic group, first trial per patient-hand; boxes are the median and IQR with whiskers at 1.5 × IQR and every observation plotted. Brackets mark pairwise Mann-Whitney comparisons surviving Holm correction within each outcome, drawn only in panels whose Kruskal-Wallis omnibus test survives Benjamini-Hochberg correction across the ten outcomes, which suppresses pairwise annotation for GME alone. GII and GOI are on the log_10_ scale on which they are defined; the crowding at the lower bound is the floor applied to error-free trials. **(B)** GNG-C in the training and held-out validation sets on one scale; the plotted quantity is the ordinal model’s linear predictor, and *C* is the concordance index within that set (0.738 training, 0.720 validation). Healthy donors (HD) are by design assigned to both sets. **(C)** Validation discrimination for each outcome alone and for GNG-C (N = 185 observations from 96 participants), with 95% percentile intervals from 2,000 bootstrap resamples of validation participants. Color and shape distinguish timing from accuracy measures: the five best single predictors are all measures of response speed. The black diamond is GNG-C, offered eight predictors and retaining five; five predictors do not exceed one. Abbreviations: criterion, response bias; d′, sensitivity; exG-μ, ex-Gaussian μ; exG-σ, ex-Gaussian σ; HD, healthy donor; iSMPT, individualized sensory-motor processing threshold; IQR, interquartile range; MS, multiple sclerosis; N, number of patient-hand observations; P-MS, progressive multiple sclerosis (SP-MS and PP-MS pooled); PP-MS, primary progressive multiple sclerosis; RR-MS, relapsing-remitting multiple sclerosis; SP-MS, secondary progressive multiple sclerosis.

However, IPS overwhelmingly drove this sensitivity. When we ranked the outcomes by effect size for the contrast between HD and P-MS, the top four metrics were all response latencies or distributional variability parameters: GRT (r = 0.648), exG-μ (r = 0.622), GRTa (r = 0.436), and exG-σ (r = 0.418); all effect sizes and adjusted p-values are provided in Supplementary Table S2. In contrast, the primary accuracy metric, GII, ranked fifth (r = 0.332). The five-predictor ordinal logistic composite (GNG-C) achieved a held-out concordance index of C = 0.720 (95% confidence interval [CI] 0.647–0.785) for disease staging (Figure 3B, Supplementary Results SR4). However, GNG-C offered no advantage over a single speed metric (Figure 3C): GRT alone achieved C = 0.723, and exG-μ reached C = 0.726.

This establishes that while the 50/50GNG is sensitive to MS-related cognitive dysfunction, its discriminative power lies in the IPS. As rSDMT and MST also measure IPS, we will need to compare their discriminatory powers side-by-side (see Results 3.6 below).

### 3.3 The 50/50GNG shows moderate internal consistency but poor between-session reliability

The measurement properties of the task further supported its characterization as an IPS test with distinct limitations for longitudinal monitoring. Within a single session, the task measured response timing consistently: we obtained a split-half r_SB_ = 0.798 for GRT, indicating good internal consistency (Figure 4A). In contrast, the r_SB_ for the commission-error rate was only 0.260, reflecting a floor effect where most participants made no errors (Supplementary Figure S2).

**Figure 4.**
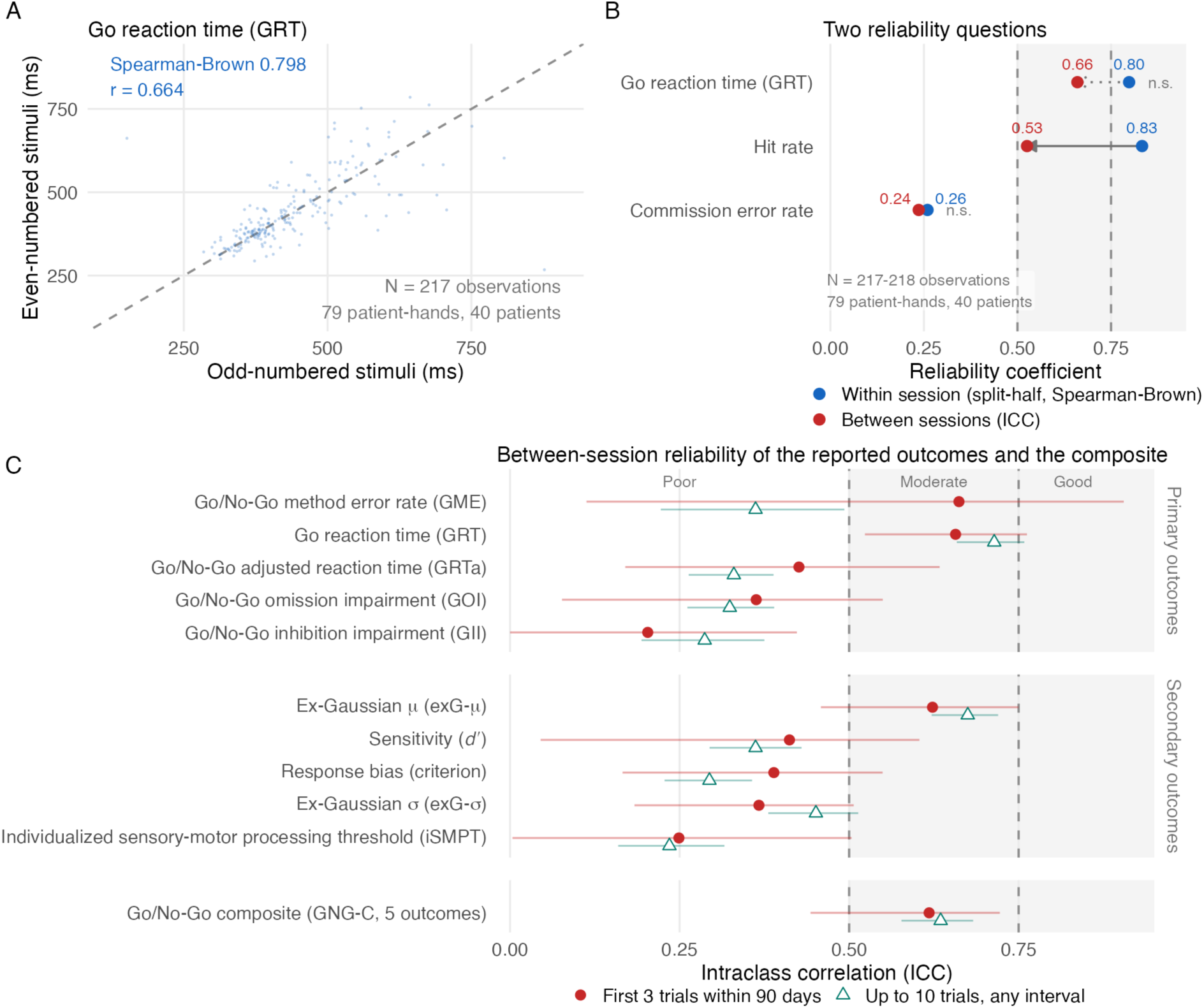
The measurement is internally consistent within a session but weakly reproducible between sessions. **(A)** Within-session split-half agreement on GRT, on the same data as panel B and panel C’s primary window: 217 observations from 79 patient-hands and 40 participants, with the median computed separately over odd- and even-numbered stimuli; splitting by alternating stimulus balances the two halves for time on task. Spearman-Brown is the split-half correlation corrected to full test length. **(B)** The comparison of the two-accuracy metrics for GRT, Hit rate and Commission error rate, each arrow running from within-session to between-session agreement. The commission-error rate sits below the other two at both ends, which is the floor rather than session-to-session change. **(C)** Between-session reliability (i.e., ICC) of the ten outcomes and GNG-C with the patient-hand as the unit: filled circles reflect the primary time window (i.e., first 3 trials within 90 days; 79 patient-hands from 40 participants), open triangles the secondary window (up to 10 trials, any interval; 381 patient-hands from 194 participants), both with 95% intervals from 2,000 patient-level bootstrap resamples. Shading marks the conventional Poor / Moderate / Good bands; no point estimate reaches Good in either window, and nine of eleven outcomes straddle 0.50 in the primary window.

However, this within-session consistency did not translate into between-session reproducibility (Figure 4B). Estimated on the data that support both coefficients (40 participants, 79 patient-hands), GRT’s agreement fell from 0.80 within session to 0.66 between sessions; the hit rate showed the same dissociation (dropping from 0.83 to 0.53), and the commission-error rate remained highly inconsistent (0.26 to 0.24). In our primary 90-day retest window, no outcome achieved good reliability (i.e., ICC > 0.75, Supplementary Results SR5). GRT was longitudinally the most reproducible outcome (ICC = 0.657 [CI: 0.523–0.762] when estimated over the full primary window rather than on the paired data shown in Figure 4B). In contrast, the primary accuracy metric, GII, performed poorly (ICC = 0.203 [CI: 0.000–0.423]). GME reached a comparable point estimate to GRT (ICC = 0.662) but with extremely broad confidence interval ([CI: 0.113–0.905]) (Figure 4C). Supplementary Table S4 reports point estimates, bootstrap CIs and sample sizes for all ten outcomes and for the GNG-C composite.

Because a reliability coefficient is a property of the cohort and its retest spacing as much as of the instrument, we benchmarked the composite against the deployed MST composite (MSC-2) on the patient-hands that contributed to both tasks’ retest windows, under a single estimator and with each composite’s published weights held fixed (Supplementary Table S5). The two were comparably reliable: GNG-C 0.670 [CI: 0.510–0.774] against MSC-2 0.692 [CI: 0.516–0.790] over the 90-day window, and 0.653 [CI: 0.594–0.702] against 0.690 [CI: 0.634–0.737] over up to ten trials.

We conclude that the observed pattern (i.e., good within-session consistency paired with poor between-session reproducibility) indicates that the 50/50GNG captures a state-dependent variable, such as daily arousal or attention state, rather than the stable biological trait required for a longitudinal biomarker. Because an already-deployed motor benchmark showed the same limitation in the same participants, this constraint is not unique to the 50/50GNG; it does mean that a single administration of either test cannot be treated as a stable individual-level measure.

### 3.4 Short-latency No-Go errors are late Go responses

By analyzing the raw event stream, we established the true origin of very short-latency No-Go errors. Across all 4,607 No-Go taps in the full extracted dataset (3,444 trials prior to trial-level quality control), 1,882 taps (40.9%) fell below the 100 ms physiological floor, and 285 exhibited negative latencies down to -569 ms, indicating that the application logged the contact before the corresponding No-Go stimulus appeared on screen. We observed identical patterns within 488 primary analysis observations: 116 sub-floor taps, 21 of which carried negative latencies.

Our subsequent analyses determined that these taps represent extremely late Go responses (beyond the response latency coded in the App design), rather than genuine failures of response inhibition. Because the application dismisses a stimulus at its 900 ms display cap regardless of whether the participant responded, a Go response slower than 900 ms lands during the subsequent blank interval (i.e., before next stimulus) or during the next stimulus, leading the software to misattribute the touch to whatever item is currently displayed (Supplementary Figure S3A).

In line with this mechanism, when we re-referenced the sub-floor taps that had followed an untapped, capped Go stimulus to that Go’s onset, all latencies fell within a plausible reaction-time window of 407 to 1,105 ms, with half clustering between 1,005 and 1,064 ms - immediately after the 900 ms cap and the nominal 100 ms blank interval (Supplementary Figure S3B). Such taps account for 72 of the 116 sub-floor taps (62.1%), a 9.4-fold enrichment over the baseline rate of 6.6% for No-Go stimuli following a capped Go (Supplementary Figure S3C). The 21 taps with negative recorded latencies arise by the same mechanism, with the finger landing inside the 100 ms blank that follows a capped Go; two such records are reproduced from the raw event stream in Supplementary Figure S4.

This logging artifact inflated commission error counts, and because the prerequisite event (untapped, slow Go responses) is itself disease-related (adjusted p-value 2.3**×**10^-4^ for P-MS versus HD), these misattributed taps were unevenly distributed across diagnostic groups. Nonetheless, diagnostic group comparisons based on commission errors remained stable regardless of attribution rules (Supplementary Results SR2, Supplementary Table S6, Supplementary Figure S3D–E).

In conclusion, this analysis identified suboptimal App design of the 50/50GNG task, which both inflated commission-error counts and left every inhibition-targeted metric null or not estimable (Supplementary Results SR6; Supplementary Figure S5), and which could be fixed in next iteration (by expanding testing time from 30 sec to e.g., 1 minute and by expanding 900 ms cap on Go stimulus to 1,200 ms), if the remaining digital biomarkers from 50/50GNG provided non-redundant clinical value to existing NeuFun-TS cognitive tests.

### 3.5 Clinical correlations of 50/50GNG digital biomarkers track global disease burden but are weaker than the Motor Sequencing Test

We observed that 50/50GNG outcomes correlated significantly with clinical and imaging measures (Figure 5A), confirming their biological validity. In MS participants, GNG-C exhibited moderate correlations with global disability scores such as CombiWISE (ρ = 0.410) and with cognitive performance measured by rSDMT (ρ = -0.534). However, these associations reflected a single, generalized disease severity factor rather than domain-specific functional localization. For example, GNG-C correlated more strongly with a whole-body neurological disability score (NeurEx total, ρ = 0.369) than with its cognitive subdomain (ρ = 0.317), demonstrating that the task measures systemic disease burden rather than isolating cognitive deficit (Figure 5A). We observed the same for correlations with imaging outcomes.

**Figure 5.**
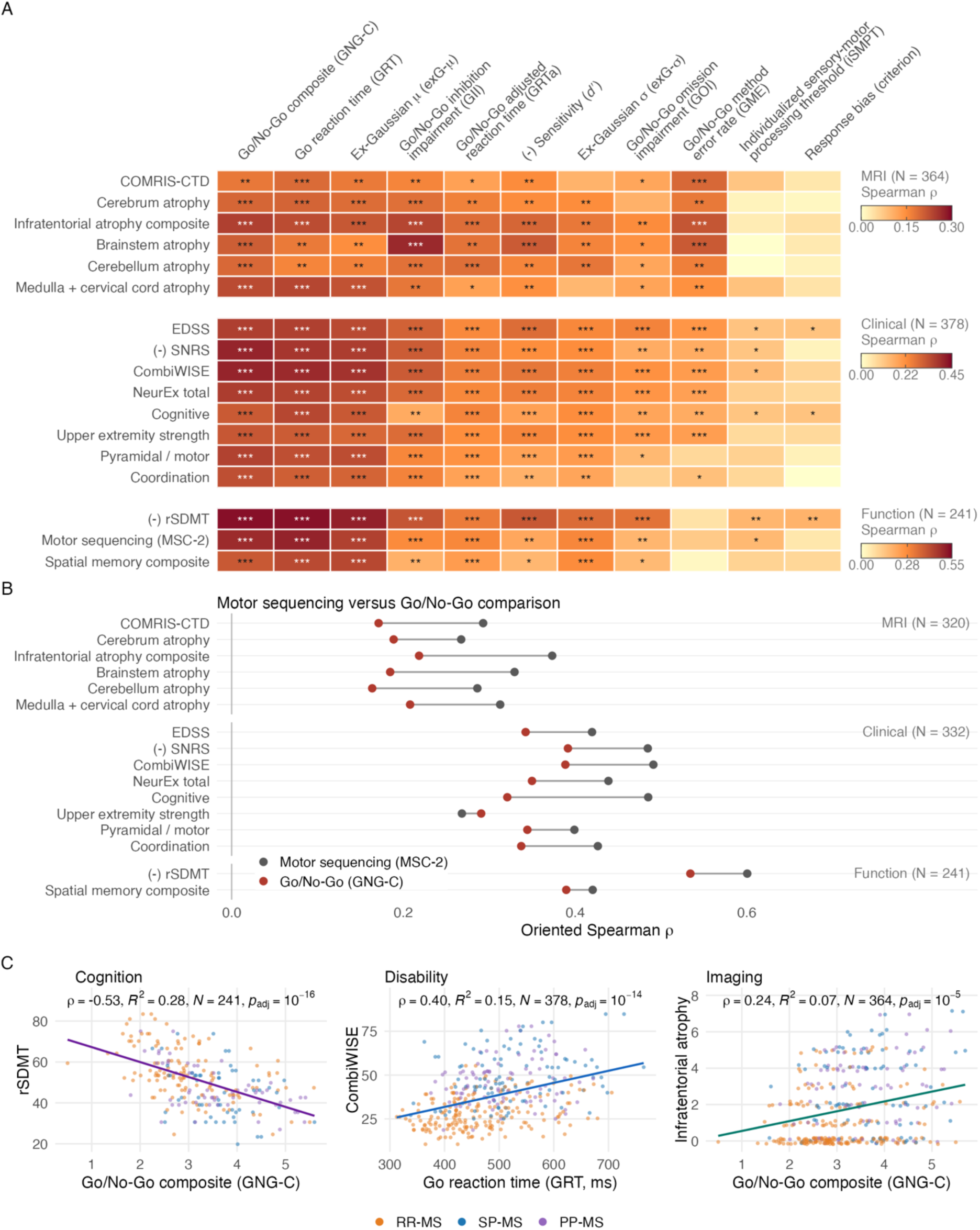
The 50/50GNG outcomes track global MS progression, and motor sequencing MSC-2 composite tracks it better in the same patients. MS patients only. **(A)** Spearman correlations between the GNG-C and the ten 50/50GNG digital biomarkers (columns) and 17 imaging, clinical and functional measures (rows), in three blocks (i.e., MRI, Clinical, other NeuFun-TS neurological functions) with their own N. Every coefficient is oriented so that a positive value means worse task performance accompanies worse disease; the rows and column that had to be sign-flipped are prefixed “(−)”.Columns are ordered by mean oriented ρ across the 17 measures drawn. The single order is imposed on all three blocks. Each block carries its own color scale, running from pale yellow at zero to dark red at that block’s strongest coefficient, so color is comparable within a block but not between blocks. Stars give the level of significance after Benjamini-Hochberg correction across all cells in the block (*** adjusted p < 0.001, ** < 0.01, * < 0.05). Of the 187 cells drawn, 33 carry no star; an unmarked cell did not reach adjusted p < 0.05 and so reads as a null rather than as an untested comparison. The four imaging rows not drawn are the regional lesion loads, in Supplementary Table S7. **(B)** The head-to-head comparison paired on the same patients: for each outcome, the correlation achieved by GNG-C and by MSC-2 from NeuFun-TS Motor Sequencing Test on the same rows. Each dumbbell is one comparative outcome; MSC-2 is higher on 18 of 20. **(C)** Three scatter plots: the strongest correlation the task produces (GNG-C against rSDMT), the best single outcome against global disability (GRT against CombiWISE), and a representative imaging cell (GNG-C against the infratentorial atrophy composite). Each panel gives Spearman ρ , its adjusted p, N and the R^2^ of the drawn line. Abbreviations: CombiWISE, Combinatorial Weight-Adjusted Disability Scale; COMRIS-CTD, Combinatorial MRI Scale of CNS Tissue Destruction; EDSS, Expanded Disability Status Scale; MRI, magnetic resonance imaging; MSC-2, two-predictor motor sequencing composite; N, number of participants; NeurEx, Neurological Exam Score; PP-MS, primary progressive multiple sclerosis; RR-MS, relapsing-remitting multiple sclerosis; rSDMT, randomized Symbol Digit Modalities Test; SNRS, Scripps Neurological Rating Scale; SP-MS, secondary progressive multiple sclerosis.

Most importantly, in direct head-to-head evaluation on identical paired participants, the motor sequencing composite (MSC-2) correlated stronger with 18 of the 20 paired clinical, functional, and imaging endpoints (i.e., the 21 pre-selected comparators less the MSC-2 itself; Supplementary Results SR7). These included 15 of the 16 measures shown in Figure 5B and three of the four regional lesion loads omitted from the figure (Supplementary Table S7). The only two exceptions were ties where both composites showed weak correlations differing by less than 0.025: upper-extremity strength (ρ = 0.291 for GNG-C vs. 0.268 for MSC-2) and cerebellar lesion volume (ρ = 0.061 for both).

### 3.6 NeuFun-TS optimization analysis: The 50/50GNG provides no meaningful added value

The final, decisive analysis confirmed that the 50/50GNG did not warrant inclusion in the optimized NeuFun-TS. While its timing measures were correlated with those of the MST (GRT vs. MST median RT, ρ = 0.506; Figure 6A), in aggregate 50/50GNG biomarkers failed to add meaningful clinical information.

**Figure 6.**
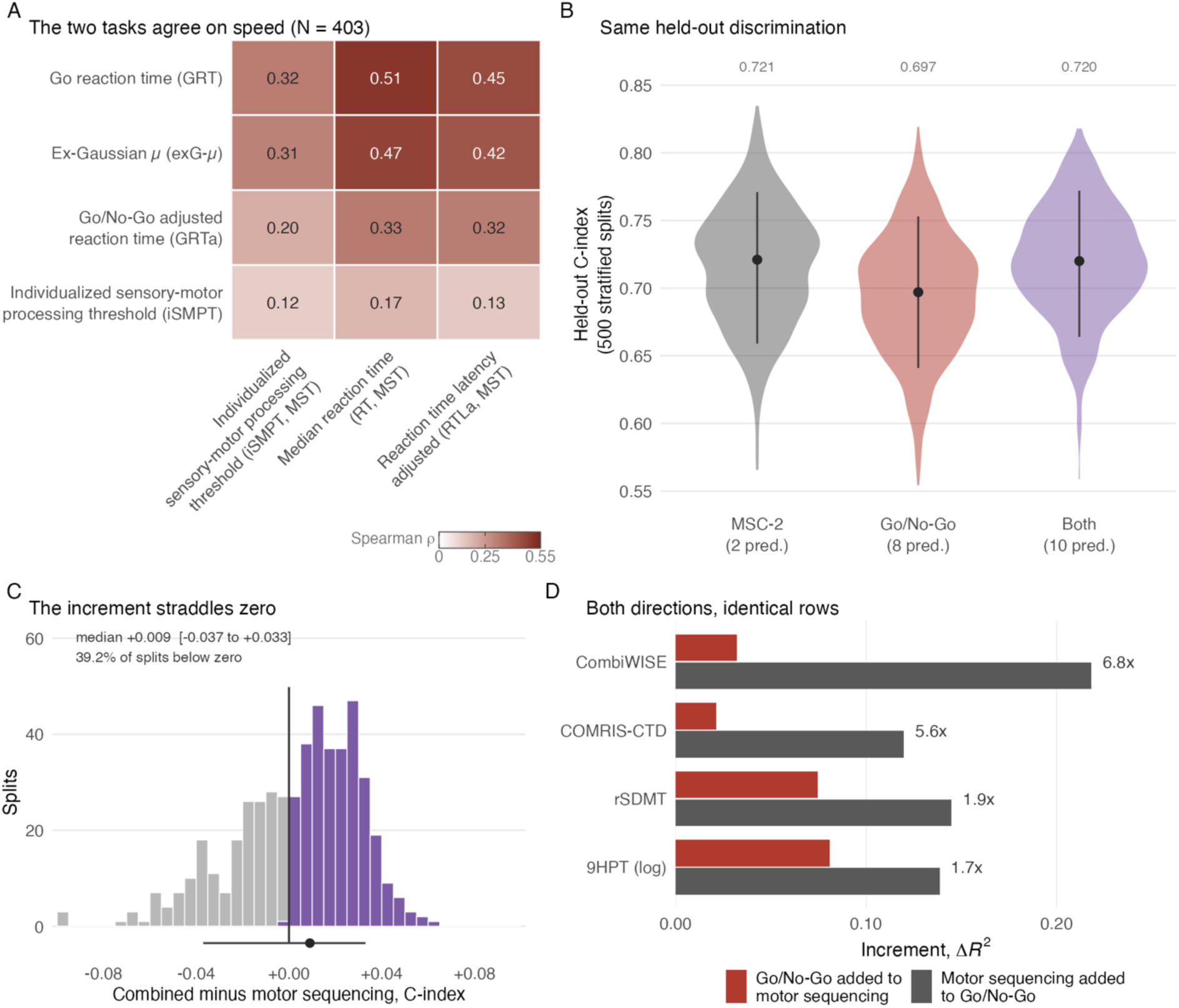
Go/No-Go biomarkers provide no added clinical value over NeuFun-TS existing cognitive tests. All panels are drawn on the 424 paired patient-hands from 217 participants (HD+MS); each panel uses the complete-case subset its measures allow (A, N = 403; B–C, 403 observations from 214 participants; D, N = 348–368). **(A)** The two tasks’ timing measures (four 50/50GNG as rows and three NeuFun-TS Motor Sequencing Test [MST] as columns) against one another, all twelve pairs, with the coefficient printed in each cell. All twelve pairs survive multiple testing correction. **(B)** Held-out concordance for three staging models (i.e., HD<RR-MS<P-MS) over the same 500 stratified splits, drawn as distributions; the median C-index is printed above each distribution and the predictor count beneath it on the axis, showing that eight GNG biomarkers do not outperform NeuFun-TS MST composite (MSC-2) either alone, or when added to MSC-2. All three models are refit within every training set. **(C)** The same 500 splits as the paired within-split difference, which provides information not available in panel B; the incremental value window of adding GNG biomarkers to MSC-2 alone straddles zero with 39.2% of splits below zero. **(D)** The same nested comparison run in both directions on identical data, for the best-performing Go/No-Go biomarker per clinical outcome. For each outcome, we are comparing added value provided by best-performing GNG biomarker against Motor sequencing biomarker alone (red bars) versus added value of Motor sequencing biomarkers when added to that same best-performing Go/No-Go biomarker (gray bars). Rows are ordered by asymmetry, which places the two global burden scales above the two direct performance tests. Abbreviations: 9HPT, 9-hole peg test; ΔR², the increment in explained variance; C-index, concordance index; CombiWISE, Combinatorial Weight-Adjusted Disability Scale; COMRIS-CTD, Combinatorial MRI Scale of CNS Tissue Destruction; HD, healthy donor; MST, NeuFun-TS Motor Sequencing Test; MS, multiple sclerosis; MSC-2, two-predictor motor sequencing composite; N, number of paired patient-hand observations; RR-MS, relapsing-remitting multiple sclerosis; rSDMT, randomized Symbol Digit Modalities Test; P-MS, progressive multiple sclerosis.

We found that adding eight 50/50GNG predictors to the baseline MSC-2 model failed to improve disease staging. The median increment in held-out concordance was only +0.009 (Supplementary Results SR8), with a 10th–90th percentile range (−0.037 to +0.033) straddling zero and yielding negative increments in 39.2% of the 500 cross-validation splits (Figure 6B-C; Supplementary Tables S8–S10). Thus, despite significantly improving in-sample fit (likelihood-ratio test p = 0.0014), adding the eight 50/50GNG parameters provides no out-of-sample diagnostic utility over MSC-2.

Because a one-directional increment cannot distinguish independent signal from two imperfect instruments failing to fully absorb one another, we ran nested linear models in both directions on identical data, comparing the unique variance each task added over the other. The contribution was highly asymmetric. Across all four outcomes (i.e., CombiWISE, the rSDMT, COMRIS-CTD and the 9HPT) the MST explained 1.7-fold to 6.8-fold more unique variance than 50/50GNG added back, and none favored 50/50GNG (Figure 6D).

These data demonstrate that although the 50/50GNG detects disease-related dysfunction, it is less discriminative and less reliable than the MST. Because its modest unique variance yields no generalization gain in disease staging, deploying 50/50GNG is not justified by its time-to-benefit ratio.

## 4. Discussion

Developing remote, patient-autonomous digital test suites requires rigorous optimization. Digital tests must not only demonstrate biological validity against established clinical and imaging benchmarks, but they must also provide sufficient incremental value to justify patient burden. In this study, we evaluated a smartphone-based sustained attention and choice-reaction task in a deeply phenotyped MS cohort. Although the task captured MS-related IPS deficits with high sensitivity, our time-to-benefit and redundancy analyses demonstrated that it adds insufficient clinical value over existing assessments to justify inclusion in NeuFun-TS.

Retaining full timestamped event streams, rather than compressing performances into summary scores proved essential for identifying suboptimal app design, where correct responses with latency exceeding 900ms coded limit were incorrectly attributed to subsequent event (i.e., either Go or No-Go) and thus mis-interpreted. This finding establishes a critical methodological caution: fixed attribution rules can silently convert processing-speed delays into apparent inhibitory errors whenever a fixed display cap overlaps the slow tail of a cohort’s reaction-time distribution. Platforms that discard raw event streams cannot detect or correct this distortion. While we could easily correct this suboptimal design in 50/50GNG by increasing time to “listen” for Go response to e.g., 1,200 ms, we decided to exclude this test from NeuFun-TS based on head-to-head comparisons with other NeuFun-TS cognitive tests^7,9,10^ with stronger psychometric properties.

Although choice-reaction performance correlated significantly with disability and MRI metrics, the MST^10^ consistently outperformed it across out-of-sample disease staging, test-retest reliability, and clinical correlations. Because NeuFun-TS already evaluates cognitive IPS via rSDMT^7^ and error monitoring through a digital spatial memory test^9^, adding 50/50GNG would lengthen autonomous testing without providing meaningful clinical gain.

The primary limitation of our study is that adopting an equal (50/50) stimulus split evaluates sustained attention and choice reaction time rather than prepotent motor restraint, limiting direct comparisons with traditional Go/No-Go literature. However, because MS-related cognitive dysfunction centers on IPS^17,18,20^, we hypothesized that a balanced paradigm would prove more sensitive in detecting deficits across MS stages, a hypothesis supported by our findings. Practical considerations also favored this design: a 50/50 split captured a representative distribution of events (∼15 Go and ∼15 No-Go stimuli) within a brief testing window. In contrast, an asymmetric ratio (e.g., 80/20) capable of inducing prepotent motor tendencies would require at least twice the duration to capture an adequate sample of No-Go trials.

To translate clinician-administered bedside assessments of neurological function into objective, self-administered digital tests, we developed a comprehensive battery of 17 distinct paradigms within NeuFun-TS. Recognizing that not all digitized bedside maneuvers would translate into viable smartphone metrics, this developmental risk was particularly acute in the cognitive domain, where we adapted tests such as Luria’s motor sequencing task and the Go/No-Go paradigm. Although clinical experience suggested that motor sequencing is more sensitive to MS-related deficits (B.B., unpublished observation), the digital implementation of the MST carried technical uncertainty regarding the algorithmic classification of finger-drawn shapes on a touchscreen. Remarkably, the digital MST outperformed the 50/50 Go/No-Go task across all evaluation criteria, even when contrasting multiple Go/No-Go metrics against a single composite MST-2 score, notwithstanding the additional, non-overlapping digital biomarkers extracted by the MST pipeline^10^.

Nonetheless, a digital Go/No-Go task may demonstrate greater utility in neurological conditions characterized by prominent frontostriatal disinhibition, such as behavioral-variant frontotemporal dementia or Huntington’s disease. For such indications, we recommend adapting the paradigm to an 80/20 Go/No-Go ratio, extending trial duration to 90–120 seconds, and lengthening the response window to at least 1,200 ms to optimize the capture and characterization of true commission errors.

Digital biomarker literature remains heavily biased toward positive findings, often obscuring redundant or suboptimal instruments. By analyzing 3,270 quality-controlled sessions from 303 longitudinally tracked participants, we demonstrate that despite high sensitivity for identifying disease-related IPS slowing and high statistical significance in disease staging, 50/50GNG does not justify deployment in NeuFun-TS alongside existing motor-cognitive tests that set very high standard for tests’ psychometric properties. Transparently reporting pruning decisions provides an empirical framework for digital test-suite optimization and prevents redundant development efforts across the field.

## Data Availability

All data produced in the present study will be available online upon acceptance of the manuscript by a peer-reviewed journal.

https://github.com/Bielekova-Lab/neufun-go-no-go

## Abbreviations

50/50GNG: Equiprobable Go/No-Go; the continuous vigilance and choice-reaction task
9HPT: 9-hole peg test
ΔR²: the increment in explained variance
C: concordance index
CI: confidence interval
CIS: clinically isolated syndrome
CNS: central nervous system
CombiWISE: Combinatorial Weight-Adjusted Disability Scale
COMRIS: Combinatorial MRI Scale
COMRIS-CTD: Combinatorial MRI Scale of CNS tissue destruction
criterion: response bias (signal detection)
d′: D-prime; sensitivity (signal detection)
EDSS: Expanded Disability Status Scale
exG-μ: ex-Gaussian μ
exG-σ: ex-Gaussian σ
exG-τ: ex-Gaussian τ
FDR: false-discovery rate
GCEd: Go/No-Go commission-error reaction-time difference
GIF: Go/No-Go inhibitory fatigue
GII: Go/No-Go inhibition impairment
GME: Go/No-Go method error rate
GNG-C: Go/No-Go composite the five-predictor ordinal composite of this study
GOI: Go/No-Go omission impairment
GPC: Go/No-Go prepotency cost
GRT: Go reaction time
GRTa: Go/No-Go adjusted reaction time
GVD: Go/No-Go vigilance decrement
HD: healthy donor
ICC: intraclass correlation coefficient
IPS: information processing speed
IQR: interquartile range
iSMPT: individualized sensory-motor processing threshold
MRI: magnetic resonance imaging
MS: multiple sclerosis
MSC-2: two-predictor motor sequencing composite (of the MST)
MST: Motor Sequencing Test
NeuFun-TS: Neurological Functions Test Suite
NeurEx: continuous disability scale computed by the NeurEx App
NIH: National Institutes of Health
NIND: non-inflammatory neurological disease
OIND: other inflammatory neurological disease
PES: post-error slowing
P-MS: progressive multiple sclerosis (SP-MS and PP-MS combined)
PP-MS: primary progressive multiple sclerosis
QC: quality control
RIS: radiologically isolated syndrome
RR-MS: relapsing-remitting multiple sclerosis
RT: reaction time
rSDMT: randomized Symbol Digit Modalities Test
SNRS: Scripps Neurological Rating Scale
SP-MS: secondary progressive multiple sclerosis

## Supplementary Material

The Supplementary Material for this article comprises Supplementary Methods SM1–SM7, Supplementary Results SR1–SR8, Supplementary Figures S1–S5 and Supplementary Tables S1–S15.

## Data and code availability

All data and code used to generate the results in this manuscript will be publicly available in the project repository at https://github.com/Bielekova-Lab/neufun-go-no-go upon publication.

## Acknowledgments

We thank our patient care coordinator Michelle Woodland for her excellent care of our patients and our former postbaccalaureate fellows for help with data collection. We thank all research participants and patients’ caregivers for their time and effort.

