## Supplementary info for "Processing Speed Accounts for the Discriminative Power of a Smartphone Go/No-Go Task in Multiple Sclerosis"

#### Supplementary Methods

##### SM1. Stimulus Sequence Generation and Empirical Characterization

We characterized the stimulus sequence directly from the recorded event data rather than relying on the application's nominal settings. We evaluated four properties: aggregate Go and No-Go proportions, variance in stimulus ratios across trials, trial-level balance, and sequential structure, including consecutive stimulus runs and transition probabilities between consecutive items.

To assess sequence randomness accurately, we applied two specific statistical baselines. 1. We benchmarked run-length structure against 3,000 simulated random shuffles of a balanced stimulus set, which tests whether stimulus order was random given its fixed composition. 2. We evaluated transition probabilities against the theoretical probability of sampling without replacement, $P(\text{Go} \mid \text{preceding Go}) = \frac{D/2-1}{D-1}$ for a balanced deck of size $D$, rather than against an uncorrected 0.5, which falsely makes any balanced finite deck look alternating.

Full empirical findings are detailed in Supplementary Results SR3 and tabulated in Supplementary Tables S11 and S12.

##### SM2. Quality Control (QC): Extended Criteria and Attribution Rules

We required each trial to contain at least one Go stimulus, one No-Go stimulus, and at least one valid Go reaction time. The balanced 30-stimulus set automatically satisfied the first two requirements; the third requirement led us to exclude nine trials, which we reviewed individually in Supplementary Results SR2 and which are counted within the 140 trial-level exclusions reported there rather than removed at a prior step.

Because the trial design makes the expected duration and stimulus count of a completed run narrow and predictable, we applied four pre-specified technical-validity criteria. First, for completeness, we excluded trials with fewer than 30 stimuli as sessions cut short. Second, for duration, we applied bounds of 28–40 s (approximately ±5 standard deviations [SD] of the raw duration distribution) to exclude mechanically interrupted or corrupted recordings. Third, for pacing, we excluded trials if the mean wall-clock time per stimulus fell below 500 ms. Fourth, for response validity, we excluded trials if more than 50% of recorded taps preceded the 100 ms physiological floor.

Finally, we applied a gate for task engagement to every trial that passed those criteria, excluding trials whose responses carried no information about the stimulus. We required $d' > 0$ and a false-alarm rate of at most 0.5. Because $d' = z(\text{hit rate}) - z(\text{false-alarm rate})$ is positive exactly when the hit rate exceeds the false-alarm rate, the first criterion cannot be failed by impairment: genuine motor or attentional impairment produces missed Go stimuli and withheld No-Go taps together, which leaves d' positive, so severe under-responding survives the gate. A non-positive d' means instead that the participant responded at least as often to the stimulus requiring withholding as to the one requiring a response, which no degree of slowness or weakness produces. The second criterion is needed because tapping at nearly every stimulus also leaves d' positive – a participant who taps without discriminating one circle from two reaches a hit rate of 1.00 with a false-alarm rate of 0.94, and so $d' = +0.64$, and a false-alarm rate above 0.5 is worse than responding at random. Both criteria are therefore properties of the response pattern rather than of speed. The gate removed 34 of the 3,304 trials that passed trial-level quality control (1.0%) and cost no participant all their data; Supplementary Results SR2 gives the counts and an illustrative case.

We applied three commission-error attribution rules to the same 488 first-trial observations to evaluate the impact of response misattribution: the raw event stream as recorded; the primary narrow rule, where we applied the 100 ms floor to No-Go taps; and a strict rule, where we additionally discarded any No-Go tap whose preceding Go stimulus timed out untapped at the 900 ms cap. We detail the results of this comparison in Supplementary Results SR2 and Supplementary Table S6.

#### SM3. Outcome Derivation: Definitions, the Commission-Error Floor and the Candidate Screen

We defined reaction time (RT) as the latency from stimulus onset to the first screen interaction. We counted a Go reaction time as valid if the participant tapped the Go stimulus and the latency fell between 100 and 2,000 ms; we excluded faster responses as anticipations or touch artifacts and slower responses as inattention rather than motor performance. 1,882 of the 4,607 No-Go taps in the raw event stream (40.9%) fell below this floor, and 285 of the 4,607 (6.2%), all of them within that sub-floor set, had negative latencies. The majority of the sub-floor taps were slow Go responses misattributed across a stimulus boundary, rather than true failures of inhibition; Supplementary Results SR6 gives the evidence.

We classified responses into four categories: hits (tapped Go stimuli), omissions (untapped Go stimuli), commission errors (tapped No-Go stimuli at or above the 100 ms floor), and correct rejections (untapped No-Go stimuli). We calculated the hit rate as hits divided by presented Go stimuli, the commission-error rate as commission errors divided by presented No-Go stimuli, and the omission rate as one minus the hit rate. Before applying logarithmic or probit transformations, we corrected extreme rates of 0 and 1 using the Hautus method[1], replacing 0 with 0.5/n and 1 with (n-0.5)/n, where n is the corresponding number of Go or No-Go stimuli. For outcomes assessing within-trial slopes, we divided each trial's stimulus sequence into four equal quarters by stimulus position rather than elapsed time.

Primary outcomes.

- GRT (Go reaction time): the median of the valid Go reaction times in the trial.
- GRTa (Go/No-Go adjusted reaction time): GRT minus iSMPT (individualized sensory-motor processing threshold; which is the median valid Go reaction time minus the fastest valid Go reaction time in the same trial – this effectively adjusts reaction time for motoric disability).
- GII (Go/No-Go inhibition impairment): $\log_{10}$ of the commission-error rate.
- GOI (Go/No-Go omission impairment): $\log_{10}$ of the omission rate, that is, of one minus the hit rate.
- GME (Go/No-Go method error rate): Go taps registering a finger count other than the required two, divided by hits: (one-finger + three-or-more-finger Go taps) / hits.

Some secondary and tertiary outcomes are based on ex-Gaussian distribution, which models reaction times (RT) by convolving a normal (Gaussian) distribution with an exponential distribution, because standard RT distributions are positively skewed (i.e., participants have a cluster of typical responses, followed by an elongated right-side tail, representing unusually slow responses). Ex-Gaussian distributions have following core parameters: μ (mu; describes center of Gaussian distribution, which is cognitively interpreted as baseline speed of sensory-motor processing); σ (sigma; described width of Gaussian distribution, which equals to standard deviation of the normal curve and cognitively it corresponds to the trial-to-trial variability in standard responses) and finally, τ (tau, which describes length and thickness of the exponential right tail and cognitively corresponds to the frequency and severity of attentional lapses). All three ex-Gaussian parameters are defined only where a trial supplies at least 10 valid Go reaction times.

Secondary outcomes.

- exG-μ (ex-Gaussian μ): the Gaussian mean of an ex-Gaussian fit to the trial's valid Go reaction times by the method of moments; $\mu = \text{mean} - \tau$, where τ is exponential component of the ex-Gaussian fit.
- exG-σ (ex-Gaussian σ): the Gaussian standard deviation of the same fit; $\sigma = \sqrt{(\text{variance} - \tau^2)}$.

- d' (sensitivity): z(hit rate) − z(commission-error rate), where z is the standard normal quantile function.
- criterion (response bias): −0.5 × [z(hit rate) + z(commission-error rate)].
- iSMPT – the minimum of the valid Go reaction times in the trial.

Exploratory outcomes.

- exG-τ (ex-Gaussian τ): the exponential component of the same ex-Gaussian fit; $\tau = (m_3 / 2)^{1/3}$, where $m_3$ is the third central moment of the valid Go reaction times, taken as 0 when $m_3 \leq 0$.
- GVD (Go/No-Go vigilance decrement) – the ordinary least squares (OLS) slope of the median valid Go reaction time on within-trial quarter, requiring at least three of the four quarters to supply a valid Go reaction time.
- GIF (Go/No-Go inhibitory fatigue) – the OLS slope of the commission-error rate on within-trial quarter, requiring at least three quarters with at least two No-Go stimuli each and a rate that is not constant across them.
- GPC (Go/No-Go prepotency cost) – the commission-error rate on No-Go stimuli preceded by a run of two or more consecutive Go stimuli, minus the rate on No-Go stimuli immediately preceded by a No-Go, requiring at least two No-Go stimuli in each of the two run classes.

Table 1 provides additional details for each of the sixteen outcomes. We highlight two impairment directions here because they invert standard interpretations: GII and GOI are negative quantities where values closer to zero indicate greater impairment, whereas d' decreases with greater impairment.

We subjected commission errors to the same 100 ms physiological floor as Go responses. The raw event stream had originally applied this floor only to Go taps. Applying the 100 ms floor provides a mechanism-agnostic correction for what we identified as a flaw in App design, where short-latency No-Go errors represent Go responses that exceed App-codified 900 ms cap (see Results 3.4). We also evaluated a stricter rule that discards any No-Go tap following a capped Go stimulus as a sensitivity analysis (Supplementary Table S6), because 23.8% of above-floor commission errors also follow a timed-out Go stimulus and cannot be separated from genuine inhibition failures on timing alone.

We screened candidate outcomes based entirely on their intrinsic measurement properties rather than their diagnostic associations. Specifically, we evaluated: estimability at this trial length (approximately 19 Go and 19 No-Go stimuli), coverage (the proportion of observations for which an outcome can be computed), within- and between-session reliability, and redundancy with existing metrics in the reported family. Six candidates fell outside the reported family based on whether we could reliably measure the construct. We classified four as exploratory (exG-τ, GVD, GIF, and GPC) and computed each on the available subset of observations. We classified two as non-estimable: PES (post-error slowing) and GCEd (Go/No-Go commission-error reaction-time difference), because each requires the occurrence of a commission error, restricting computation to a non-representative, performance-selected subset. We excluded all six from the multiple-testing denominator but retained and analyzed them in full in Supplementary Results SR6. We assessed reliability across all 16 candidates and summarized the complete screening results, including every reliability coefficient, in Supplementary Table S13.

#### SM4. Multivariable Staging: Protocol, Diagnostics and Sensitivity

We split the multiple sclerosis (MS) cohort 70/30 into training and validation sets, stratifying on diagnosis, sex, age tertile and NeurEx (digitalized neurological examination) total score and partitioning by participant so that no MS participant contributed to both sets; we assigned healthy donor (HD) participants to both sets. The model orders severity in three levels: HD < relapsing-remitting MS (RR-MS) < progressive MS (P-MS), the latter comprising the secondary progressive (SP-MS) and primary progressive (PP-MS) subtypes. We performed stepwise Akaike Information Criterion (AIC) selection inside the training set only and evaluated the selected model once in the held-out set, reporting discrimination as the validation concordance index (C). We assessed selection stability by rerunning the whole protocol 100 times with a different split each time, and we report a further sensitivity analysis dividing the HD participants 70/30 in Supplementary Results SR4.

#### SM5. Reliability: Estimator, Windows and Robustness

We estimated reliability as the two-way consistency, single-measures intraclass correlation (ICC) of Shrout and Fleiss[2], taking the patient-hand as the unit of analysis and the repeated session as the second factor. Consistency rather than absolute agreement means that a systematic shift between sessions (e.g., due to MS progression) is fitted and excluded from the error term. We obtained the coefficient from an equivalent linear mixed model with random intercepts for patient-hand and session.

We defined three retest windows. The primary window took the first three trials within 90 days; the secondary window took up to ten trials per patient-hand with no interval restriction; and as an exploratory check we computed a third, unrestricted window over the entire follow-up with no cap. Supplementary Results SR5 reports the coefficients from all three.

We report 95% percentile confidence intervals (CI) from 2,000 bootstrap resamples in which we resampled participants rather than observations.

We calculated the reliability of the Go/No-Go composite (GNG-C) by applying the fixed training-set coefficients to every retest session, which reflects the reliability of the rule a deployed composite would use.

#### SM6. Cross-Task Comparison: Full Specification and Protocol Sensitivity

We paired records of the Motor Sequencing Test (MST) and the equiprobable Go/No-Go task (50/50GNG) within participant and hand, keeping the temporally closest pair of sessions within ±7 days. We refit the motor sequencing composite (MSC-2) on this overlap, retaining its two published predictors[3] but re-estimating all coefficients, so that the comparison against the Go/No-Go model was fair.

We tested incremental diagnostic staging with ordinal logistic regression on the three-level severity ordering, comparing motor sequencing alone, Go/No-Go alone, and the two combined. We report held-out concordance as the median and 10th–90th percentile over 500 patient-level 70/30 splits.

We recomputed each Go/No-Go association with a clinical outcome with the motor sequencing speed measures adjusted for, and we report the proportion of the raw association retained.

For every combination of Go/No-Go outcome and clinical measure we compared two nested linear models: the motor sequencing predictors alone, and those predictors plus one Go/No-Go outcome (the best GNG predictor for that outcome), which gives the increment in explained variance ($\Delta R^2$) attributable to that outcome; we tested each increment by F test with Benjamini-Hochberg correction. We computed the same increment in both directions on identical rows and report their ratio as an asymmetry.

The ordinal model of this section stratifies its splits on diagnostic group alone and divides the HD participants 70/30 like everyone else, whereas the staging model of Section 3.2 stratifies on diagnosis, sex, age tertile and NeurEx total and assigns HD participants to both sets (SM4). We varied both choices factorially (i.e., group strata or the finer Section 3.2 strata, HD participants divided or replicated) and ran each of the four resulting protocols to 200 reseeds. Supplementary Table S10 gives all four; the incremental-staging analysis of Section 3.6 reports the first row.

#### SM7. Discriminant Validity Against Non-MS Neurological Disease

We held the three non-MS groups: non-inflammatory neurological disease (NIND), other inflammatory neurological disease (OIND) and clinically or radiologically isolated syndrome (CIS/RIS) out of every primary analysis. We evaluated GNG-C on these groups with the coefficients of the model from Section 3.2 held fixed. We compared all seven groups by Kruskal-Wallis test and then compared all 21 group pairs by Wilcoxon rank-sum test with Benjamini-Hochberg correction across the full set of 21.

### Supplementary Results

#### SR1. Discriminant Validity: GNG-C Against Non-MS Neurological Disease

When we applied to 110 held-out non-MS observations, it varied significantly across all seven diagnostic categories (Kruskal-Wallis H = 115.9, $p = 1.2\times10^{-22}$). The GNG-C robustly separated HD from other inflammatory neurological diseases (OIND; median 2.32 vs 3.43; r = 0.47, $q = 1.0\times10^{-4}$) whereas non-inflammatory neurological diseases (NIND) remained indistinguishable from controls (median 2.56; r = 0.11, q = 0.36), confirming this sensitivity is not a generic artifact of non-inflammatory pathology or age. Furthermore, both non-MS disease cohorts scored substantially lower than progressive MS subtypes (e.g., NIND vs SP-MS, r = 0.62, $q < 10^{-7}$), demonstrating preserved dynamic range without premature ceiling effects. Overall, 15 of 21 pairwise contrasts remained significant after Benjamini-Hochberg false-discovery-rate adjustment. Crucially, the two planned non-significant contrasts supporting task construct specificity were HD versus NIND and CIS/RIS versus RR-MS (all group medians and 21 pairwise contrasts appear in Supplementary Tables S14 and S15).

#### SR2. Quality Control: Exclusions and Attribution-Rule Comparison

No exclusion removed a participant on the grounds of slow performance. The nine trials excluded for having no valid Go RT were reviewed individually; none reflected slow performance but rather instruction reversals or trials with no screen contact. Trial-level quality control removed 140 of 3,444 trials in total (4.1%): 131 failing one of the four technical validity criteria and the nine above lacking a valid Go reaction time. Additionally, the subject-level gate for non-discriminating responses removed another 34 of the remaining 3,304 trials (1.0%). No participant in any primary group lost all their data. One illustrative case was a HD whose first session (32/32 hits, 29/31 commission errors) registered a catastrophic GNG-C score of 5.58 (above the third quartile of progressive MS) due to misunderstanding the instructions, while nine subsequent sessions were nearly error-free; the gate's false-alarm bound removed that session, so it enters none of the reported analyses.

A three-rule comparison for commission-error attribution (raw, narrow 100 ms floor, strict) showed that while tightening the rule increased the proportion of error-free trials from 48.8% to 64.3%, the GII group separation effect size remained stable (0.326, 0.332, 0.322), demonstrating that the floor effect rather than the attribution rule, is the limiting factor (Supplementary Figure S3D–E; Supplementary Table S6). The 100 ms floor was therefore adopted on construct validity: it ensures that error rates do not count responses intended for a prior stimulus.

#### SR3. Stimulus Composition and Sequential Structure

Empirical analysis confirmed that the stimulus generator draws without replacement from a balanced 30-stimulus set. The informative finding is the under-dispersion around the aggregate Go proportion (0.4994 across 126,237 stimuli), which establishes that balance is enforced by design rather than achieved on average: every one of the 3,270 analyzable trials presented exactly 15 Go and 15 No-Go stimuli in its first 30. This fixed the minimum length of a completed trial, informed the quality-control thresholds in Supplementary method SM2. While 50/50 composition was fixed, order within the set was a free shuffle, so the stream remained unpredictable. Run-length statistics were indistinguishable from simulated shuffles of the same 15/15 set (longest Go run 4.73 vs 4.71, Kolmogorov–Smirnov [KS] $p = 1.00$; number of runs 15.99 vs 16.00, KS $p = 0.23$), and the lag-1 transition probabilities sat at the values that drawing without replacement requires ($P(\text{Go} \mid \text{Go}) = 0.4835$ against an expected 14/29 = 0.4828; $P(\text{Go} \mid \text{No-Go}) = 0.5170$ against 15/29 = 0.5172). The key interpretive consequence, as noted by Wessel[4], is that an equiprobable design does not establish a strong prepotent response tendency, framing the task as a test of stimulus discrimination under time pressure rather than classical response inhibition. (See Supplementary Tables S11 and S12.)

#### SR4. Multivariable Disease Staging (HD<RR-MS<P-MS): Diagnostics and Model Stability

Stepwise AIC selection was applied to first-trial observations with complete data on all ten reported outcomes (N = 459 / 488); eight were offered to the selection, GRT and exG-μ being withheld as collinear with the retained GRTa/iSMPT pair (GRTa = GRT − iSMPT exactly; exG-μ against GRT, $\rho = 0.95$). In an initial training set (338 observations) stepwise selection yielded a five-predictor Go/No-Go composite score (GNG-C: GOI, GRTa, GME, d', iSMPT) achieving a concordance index of C = 0.738. The concordance index remained stable in the out-of-sample validation set (185 observations), reaching C = 0.720 (95% CI 0.647–0.785).

However, cross-validation revealed that the multivariable model offers no meaningful discriminative advantage over single timing parameters (exG-μ C = 0.726, GRT C = 0.723).

Furthermore, GNG-C feature composition was highly sensitive to data partitioning: A stability analysis over 100 different 70/30 splits showed that while GRTa and iSMPT were always retained, other predictors were draw-dependent (e.g., GOI was retained in only 33% of reruns), though the median concordance remained stable at 0.739. Dividing the HD 70/30 rather than replicating them into both sets lowered the median held-out concordance over 100 splits from 0.739 to 0.706 (10th–90th percentile 0.657–0.751) and left the number of selected predictors unchanged at a median of five.

#### SR5. Reliability: Robustness Checks and Full Coefficients

The Go/No-Go paradigm demonstrated moderate within-session internal consistency for timing metrics, but between-session test-retest reproducibility was poor across nontiming domains: Split-half reliability (Spearman-Brown corrected) for GRT was adequate across both the full first-trial cohort ($r_{SB}$ = 0.777; N = 485-488 observations across 245-247 participants) and the retest cohort ($r_{SB}$ = 0.798; N = 217-218 observations across 79 patient-hands and 40 participants). In contrast, split-half reliability for the commission-error rate was uniformly poor across both samples ($r_{SB}$ = 0.326 and $r_{SB}$ = 0.260, respectively; Figure 4 and Supplementary Figure S2). This indicates that the severe floor effect on commission errors reflects an intrinsic limitation of the measure itself under an equitable 50/50GNG design, rather than sample-specific noise.

Between-session reproducibility in the primary 90-day retest window was moderate for timing features but poor for inhibitory and signal-detection metrics. Intraclass correlation coefficients (ICC) ranked as follows: GME (0.662), GRT (0.657), exG-μ (0.623), and the five-predictor composite GNG-C (0.618), followed by substantial drops in GRTa (0.426), d' (0.412), criterion (0.389), exG-σ (0.367), GOI (0.363), iSMPT (0.249), and GII (0.203). Restricting the GNG-C analysis to patient-hands matched to both tasks marginally elevated its ICC to (0.670), which remained slightly below the comparator Neurological Functions Test Suite (NeuFun-TS) Motor Sequencing Composite (MSC-2: 0.716 on its dedicated retest cohort, and 0.692 on the matched cohort; Supplementary Table S5).

Expanding the evaluation beyond the primary window confirmed that only baseline reaction-time parameters retain reproducibility over time. In the secondary window (up to ten trials per hand with no fixed time restriction), moderate reliability was preserved exclusively by GRT (ICC = 0.714) and exG-μ (ICC = 0.675). Similarly, across an unrestricted long-term follow-up (3,053 observations across 381 patient-hands and 194 participants), this exact hierarchy was preserved: exG-μ (ICC = 0.694) and GRT (ICC = 0.687) remained the most stable, followed by exG-σ (ICC = 0.500), while all remaining metrics fell at or below 0.430. Sensitivity analyses verified that these attenuated between-session coefficients reflect true lack of longitudinal reproducibility rather than artifacts of retest intervals or cohort selection.

#### SR6. The Short-latency No-Go taps are Go Responses Falling Beyond App-coded 900 ms Cap

Short-latency No-Go taps are late Go responses. In the raw event stream, 1,882 of 4,607 No-Go taps (40.9%) fell below the 100 ms physiological floor, and 285 of them had negative latencies (i.e., the application logged the tap before the stimulus had appeared). We worked out what was happening on the 488-observation primary dataset, which contains 116 of these sub-floor taps. For 72 of the 116 (62.1%), the stimulus immediately before was a Go that the participant never tapped and that timed out at its 900 ms cap. Only 6.6% of No-Go stimuli follow an untapped Go, so sub-floor taps land in that position 9.4 times more often than chance alone would produce. Each of those earlier Go stimuli ended at exactly 900 ms, so there is no ambiguity about which stimulus a late tap belonged to and no need to allow a margin around the cap. Measured from the onset of that earlier Go rather than the No-Go, all 72 latencies land in a range of plausible responses: 407 to 1,105 ms, with half between 1,005 and 1,064 ms, just past the 900 ms cap plus the 100 ms blank that follows it. These taps are therefore slow Go responses that the application incorrectly assigned to the next stimulus (Supplementary Figure S3B).

Negative latencies are caused by logging fault, not subject behavior. A negative latency means the application recorded a tap as a response to a stimulus that had not yet appeared, which is biologically implausible. Across all 4,607 No-Go taps, every tap with a negative latency also has a recorded display duration of zero, its onset and offset timestamps being identical – again, this is biologically implausible and contrasts with uniformly non-negative latencies recorded for every tap with a real duration (see example in the Supplementary Figure S4).

The No-Go stimuli in these records were classified normally, and the log shows it. From a zero-duration onset stamp to the next logged row is a median of 901 ms, the same median as No-Go records carrying a real duration, and 19 of the 21 spans match the 900 ms cap. Because a No-Go disappears as soon as it is touched (i.e., tapped records last a median of 338 ms) those 19 stimuli behaved exactly like stimuli nobody touched while they were on screen. A duration of zero therefore means the application failed to write the offset field, not that the stimulus was absent. In behavioral terms these were correct rejections, counted as commission errors only because a tap arrived just before them. We changed no analysis on account of these records, based on three reasons we'll describe here: First, the 100 ms floor already discards every one of them, so GII, the inhibition outcome we report, never sees them. Second, they stay in the No-Go denominator, where 21 of 9,361 stimuli (0.2%) cannot shift any rate; discarding stimuli that were shown is a bigger methodological step than an error of this size justifies. Third, we still score the earlier Go stimulus as an omission, because the task counts a response only if the finger lands while the stimulus is on screen, and by then it was gone. Crediting these taps as very slow hits would change what the measure means, from "responded in time" to "responded eventually", and there is no principled deadline for the latter. Scoring them as omissions is the conservative choice and matches what the deployed application reports for the trial. One consequence of our choice is that GOI and GII are no longer independent, because one slow response can produce both an omission on the Go stimulus it was meant for and a commission error on the No-Go stimulus it was charged to. We left trial-level quality control alone as well, since a trial in which more than half the taps fall below the floor is already excluded. Eighteen of the 488 observations (3.7%) contain at least one negative latency tap.

Because a 50/50 Go/No-Go design does not establish a dominant Go response, inhibitory demand failed to produce the expected effects across all target metrics (Supplementary Figure S5). The metrics intended to reflect inhibitory load yielded either null results, paradoxical directions, or severe data-coverage constraints: The prepotency cost (GPC) gave the cleanest null: rank-biserial $r = 0.001$ (95% CI −0.139 to 0.143), $p = 0.99$. The vigilance decrement (GVD) was not significant ($r = +0.119$, $p = 0.14$); likely because a 30-second trial is too short for a time-on-task effect to build. exG-$\tau$ was significant but pointed the wrong direction ($r = -0.176$, $p = 0.034$). Inhibitory fatigue (GIF) was null and poorly covered ($r = -0.047$, $p = 0.71$; defined for 215 of the 488 observations, 44.1%). Post-error slowing (PES) could be computed for only 3.1% of observations (15 of 488, one of them a HD), so we consider it not estimable. GCEd, the commission-error RT difference, gave a large effect ($r = -0.384$), but it was defined on a performance-selected subset (i.e., the same 215 observations as GIF, since both require at least one commission error) and was not robust; what it does show is that commission errors were faster than correct responses by a median of 168 ms, consistent with impulsive responding under time pressure.

#### SR7. Clinical Correlation and Head-to-Head Comparison Details

The composite score (GNG-C) exhibited the highest associations with the four global disability scales, correlating with the Combinatorial Weight-Adjusted Disability Scale (CombiWISE) at $\rho = 0.410$. However, univariate Go reaction time (GRT) performed essentially on par, falling within 0.03 correlation points of the multivariable composite across all four scales. GNG-C showed its strongest association with the NeuFun-TS randomized Symbol Digit Modalities Test (rSDMT, $\rho = -0.534$), a benchmark that GRT matched almost identically ($\rho = -0.532$).

Rather than isolating distinct cognitive or inhibitory dimensions, the correlation profile across clinical comparators reflects a shared general disability factor. Metric effect rankings remained virtually identical across clinical examination and functional testing blocks (rank correlation $\rho = 0.909$). Furthermore, 9 of the 11 Go/No-Go metrics correlated more strongly with whole-body neurological impairment (NeurEx total score) than with regional upper-extremity motor disability; for example, GRT correlated with NeurEx at $\rho = 0.358$ compared to $\rho = 0.313$ for hand-specific strength.

Finally, a direct head-to-head comparison on matched observations demonstrated the superior clinical sensitivity of the NeuFun-TS motor sequencing benchmark (MSC-2). MSC-2 yielded stronger correlations than the Go/No-Go outcomes on 18- of the 20-comparator metrics, with marked advantage on the cognitive examination panel ($\rho = 0.485$ versus 0.321).

#### SR8. Cross-Task Comparison: The Three Candidate Tests

We evaluated whether 50/50GNG digital biomarkers improve disease staging beyond the performance of NeuFun-TS motor sequencing test. Among 403 complete paired patient-hands (of 424), a refit two-predictor MSC-2 model achieved a median held-out concordance of $C = 0.721$, an eight-predictor Go/No-Go model reached $C = 0.697$, and a combined model reached $C = 0.720$. Within-split paired comparisons, which reflect true incremental utility rather than marginal median differences, showed that adding 50/50GNG biomarkers to MSC-2 yielded a negligible median gain of +0.009 (10th–90th percentile: -0.037 to +0.033), resulting in negative increments across 39.2% of splits. While the combined model improved training fit (likelihood-ratio test $p = 0.0014$), this required eight additional parameters and failed to translate out-of-sample, indicating parameter overfitting rather than genuine diagnostic value.

The two tasks share substantial explained variance. After adjusting for motor sequencing speed, Go/No-Go inhibition metrics retained on average 61.4% of their unadjusted effect sizes; although not entirely redundant, the residual signal is modest. Most importantly, incremental variance partitioning was markedly asymmetric: On the 9-hole peg test (9HPT), d' added $\Delta R^2 = 0.081$ over an MSC-2 baseline, whereas MSC-2 added 1.7-fold more variance over a 50/50GNG baseline. This asymmetry expanded to 6.8-fold on global disability scales such as CombiWISE, demonstrating that motor sequencing dominates their shared predictive variance.

This comparison was sensitive to only one methodological factor: whether HD were duplicated across training and validation partitions. When HD were strictly partitioned across sets (200 random splits per protocol; Supplementary Table S10), the median concordance increment from adding 50/50GNG remained +0.010 (36.0% negative splits). In contrast, duplicating HD across sets artificially boosted this increment to +0.030 (only 2.5% negative splits). Adding finer stratification variables had negligible impact (+0.012; 35.5% negative splits), yet produced the same artificial inflation once combined with HD duplication (+0.034; 1.5% negative splits).

The reason is straightforward: HD cohort sitting in both sets is an easy observation scored twice, which enlarges the held-out set (median 161 against 120 observations) and helps the 50/50GNG predictors most, increasing their median concordance from 0.699 to 0.736 against 0.720 to 0.726 for motor sequencing. Section 3.6 therefore adopts the strict non-overlapping partition to prevent data leakage and optimistic bias. Regardless of partitioning protocol, the incremental concordance remained < 0.05 – well within the stochastic error of random resampling (Supplementary Table S10).

### Supplementary Figures

#### Supplementary Figure S1

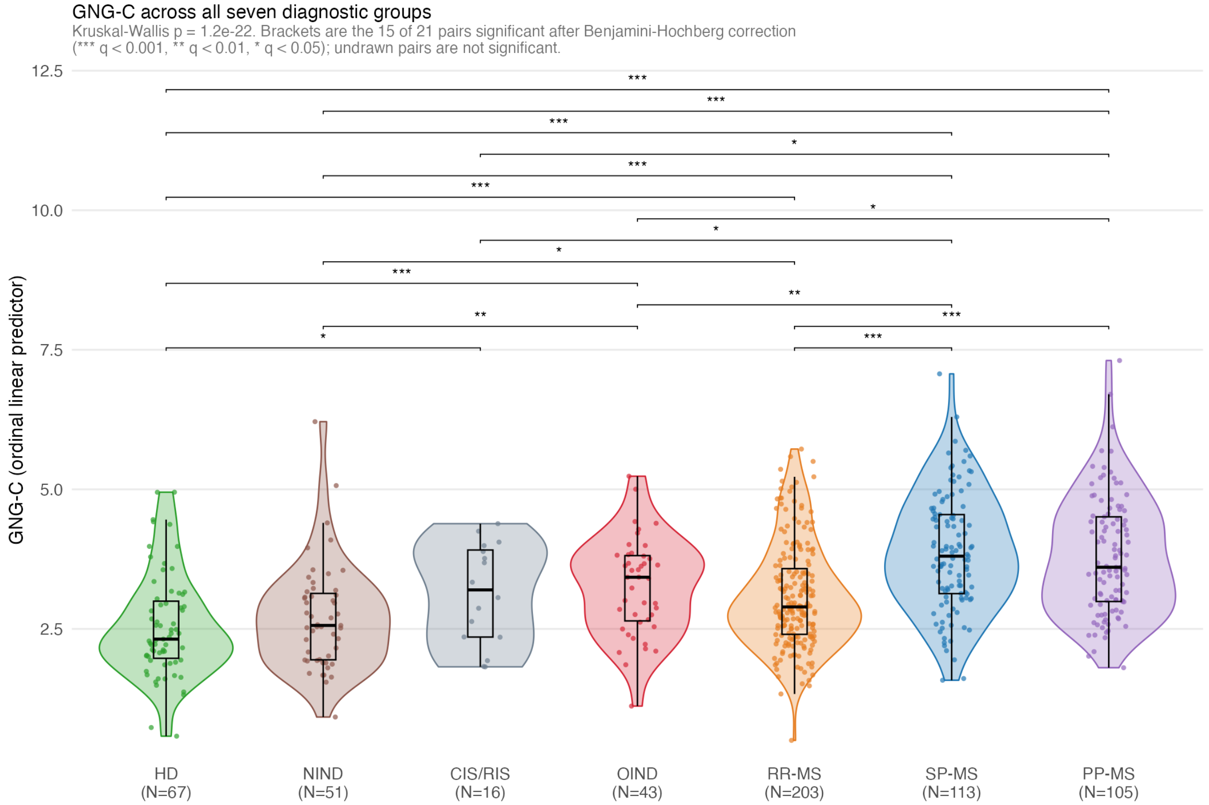


**Supplementary Figure S1. GNG-C discrimination across all seven diagnostic groups**: the four groups of the primary analysis (i.e., HD + three MS categories) together with the three non-MS groups held out of it, on first-trial patient-hand observations. Violins are trimmed to each group's own range, boxes give the median and quartiles, with individual subjects shows as dots. The three held-out non-MS groups retain the brown, strawberry red and steel gray of main-text Figure 1 and are ordered by ascending median, so the gradient reads left to right; the four HD and MS groups keep the colors and order of main-text Figure 3. Axis labels contain each group's N. Brackets are the 15 of 21 pairwise contrasts significant after Benjamini-Hochberg correction across all 21 (*** p < 0.001, ** p < 0.01, * p < 0.05); pairs without a bracket are the non-significant ones. Abbreviations: CIS/RIS, clinically or radiologically isolated syndrome; GNG-C, Go/No-Go composite; HD, healthy donor; N, number of first-trial patient-hand observations; NIND, non-inflammatory neurological disease; OIND, other inflammatory neurological disease; PP-MS, primary progressive multiple sclerosis; q, Benjamini-Hochberg-adjusted p-value; RR-MS, relapsing-remitting multiple sclerosis; SP-MS, secondary progressive multiple sclerosis.

Supplementary Figure S2

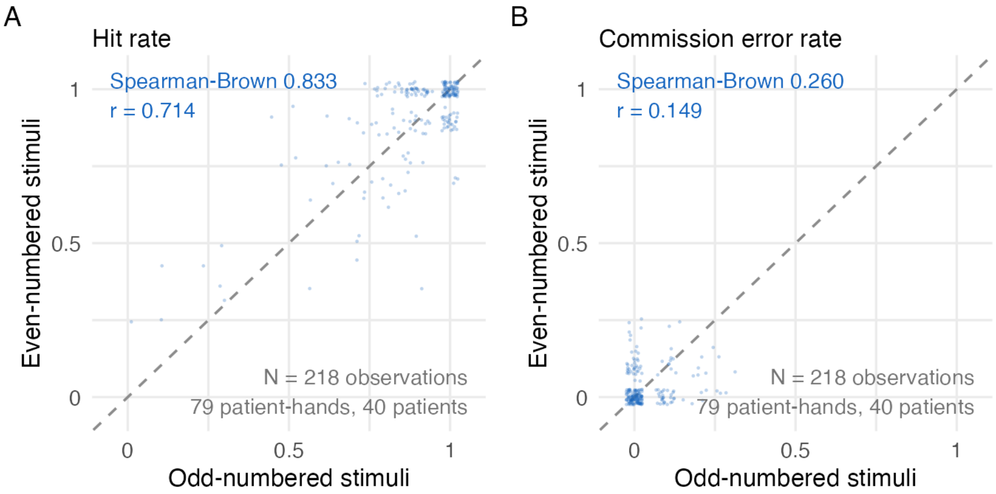


**Supplementary Figure S2. Poor within-session reliability of error-related biomarkers**. Both panels are constructed exactly as main-text Figure 4A and are drawn on the same rows as Figure 4A and 4B — the primary retest window (the first three trials per patient-hand within 90 days), 218 within-session observations from 79 patient-hands of 40 participants. **(A)** Hit rate, odd- against even-numbered stimuli ($r_{SB}$ = 0.833; uncorrected Pearson correlation = 0.714). **(B)** Commission-error rate ($r_{SB}$ = 0.260; uncorrected Pearson correlation = 0.149). Each rate is a proportion over roughly ten stimuli per half — the ~10 Go stimuli for the hit rate, the ~10 No-Go stimuli for the commission-error rate — so each takes only about eleven attainable values, spaced 0.1 apart. Points are therefore displaced by up to ±0.025 in each axis, a quarter of that spacing, so overlapping observations become visible without any point reaching a neighboring attainable value; the dashed line is the identity. The two measures pile up at opposite extremes: 77 of 218 observations have a hit rate of exactly 1.0 in both halves, and 123 of 218 (56.4%) have a commission-error rate of exactly 0 in both halves. Both coefficients therefore describe concordance across coarse, discrete bins rather than a continuous-scale linear association. Commission errors are the more severely truncated of the two: with more than half of observations at zero in both halves, too little between-participant variance remains for the two halves to covary reliably, which is the floor effect Section 3.3 and Supplementary Results SR5 report. Abbreviations: N, number of within-session observations; $r_{SB}$, the Spearman-Brown coefficient — the uncorrected Pearson correlation between the two halves corrected to full test length, 2 x (Pearson r)/(1+ Pearson r).

Supplementary Figure S3

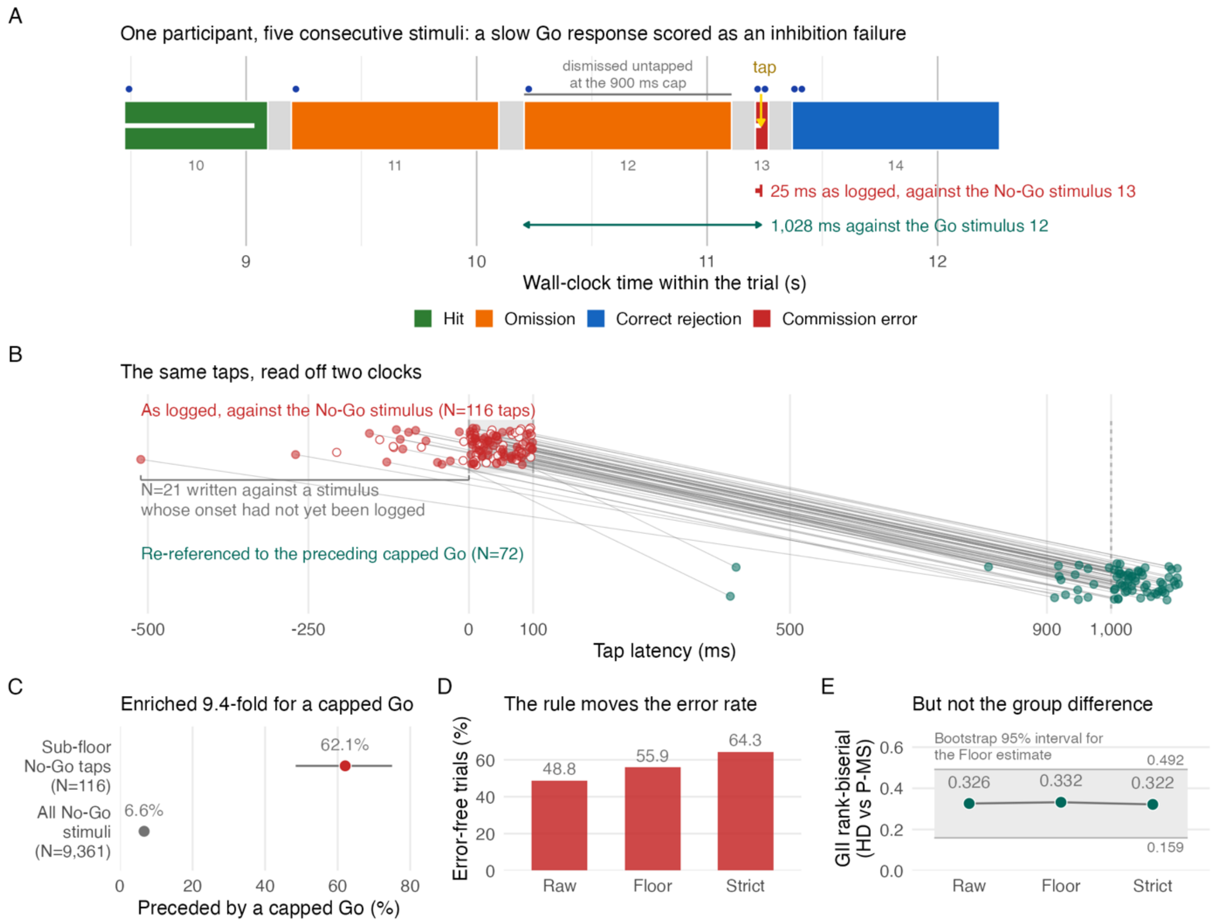


**Supplementary Figure S3. Short-latency No-Go errors are late Go responses**. **(A)** Representative sequence illustrating the artifact across five consecutive stimuli in one participant with PP-MS (baseline median Go reaction time [GRT] = 620 ms). Stimuli 11 and 12 expire at the 900 ms display cap without a response. A contact registered 25 ms after No-Go stimulus 13 appears, is logged incorrectly as a commission error; referenced to stimulus 12 onset, this contact falls at 1,028 ms, which is a typical slow Go response. The standard 100 ms post-stimulus floor successfully excludes this event from analysis. **(B)** Sub-floor No-Go taps (N = 116 across 488 observations) evaluated in App-logged versus stimulus-preceding reference frames. Gray lines connect identical events across both timelines. Top row: logged latency relative to No-Go onset, showing the 0-100 ms exclusion zone and 21 logging artifacts with negative timestamps. Bottom row: the subset immediately following an unresponded, capped Go stimulus (N = 72; filled circles), re-referenced to that prior Go onset. These events cluster tightly around the expected timeout-plus-interstimulus interval (median 1,026 ms, IQR 1,005–1,064 ms; dashed line at 1,000 ms), confirming they represent delayed Go responses. **(C)** Mechanistic enrichment: 62.1% of sub-floor No-Go taps were preceded by an unresponded Go stimulus, compared to a baseline rate of only 6.6%

across all No-Go presentations (9.4-fold enrichment; bars denote 95% patient-clustered bootstrap CIs across 2,000 resamples). **(D, E)** Sensitivity of task outcomes across three error-attribution filters: raw logs, the 100 ms floor, and a strict filter discarding any No-Go tap preceded by a timed-out Go. **(D)** Applying the filters increases the proportion of zero-commission trials from 48.8% to 64.3%. **(E)** Despite this shift in error counts, diagnostic discrimination (GII rank-biserial correlation between HD and progressive MS) remains stable ( r = 0.322–0.332), shifting across a margin 33-fold narrower than the 95% bootstrap confidence interval (0.159–0.492, shaded band). The 100 ms floor is therefore justified on construct validity rather than empirical bias correction (see Section 3.4, SR6, and Supplementary Figure S4). Abbreviations: CI, confidence interval; GII, Go/No-Go inhibition impairment; GRT, Go reaction time; HD, healthy donor; IQR, interquartile range; N, number of observations (taps, stimuli or patient-hands as stated in each panel); P-MS, progressive multiple sclerosis (SP-MS and PP-MS combined); r, rank-biserial correlation.

Supplementary Figure S4

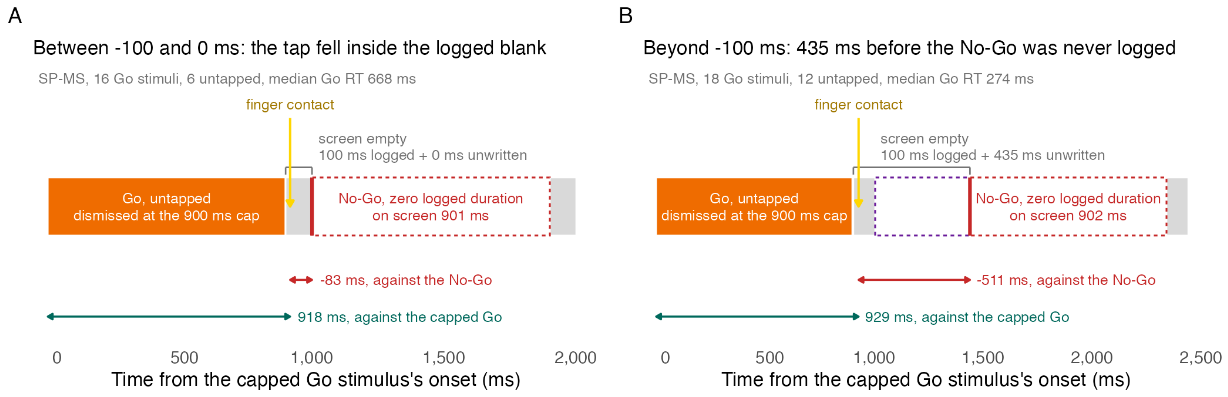


C

The rows the application wrote (ms, same origin)

| record | onset | offset | duration | tap at | contact |
|---|---|---|---|---|---|
| Go | -1,003 | -103 | 900 | no tap | |
| blank | -101 | -1 | 100 | no tap | |
| Go | 0 | 900 | 900 | no tap | |
| blank | 901 | 1,001 | 100 | no tap | |
| No-Go | 1,001 | none | 0 | -83 | 918 |
| blank | 1,902 | 2,002 | 100 | no tap | |
| No-Go | 2,004 | 2,904 | 900 | no tap | |

latency = contact - No-Go onset = 918 - 1,001 = -83 ms
-83 ms = -( 83 ms inside the logged blank + 0 ms unwritten )

D

The same fields, extreme case

| record | onset | offset | duration | tap at | contact |
|---|---|---|---|---|---|
| No-Go | -1,006 | -106 | 900 | no tap | |
| blank | -102 | -2 | 100 | no tap | |
| Go | 0 | 900 | 900 | no tap | |
| blank | 905 | 1,005 | 100 | no tap | |
| No-Go | 1,440 | none | 0 | -511 | 929 |
| blank | 2,342 | 2,442 | 100 | no tap | |
| No-Go | 2,442 | 3,342 | 900 | no tap | |

latency = contact - No-Go onset = 929 - 1,440 = -511 ms
-511 ms = -( 76 ms inside the logged blank + 435 ms unwritten )

**Supplementary Figure S4. Mechanism of negative response latencies and logging artifacts.** Drawn from raw event logs across 488 observations containing 21 No-Go taps recorded with negative latencies: **(A, C)** Latencies between -100 ms and 0 ms (N = 10). A Go stimulus timed out at its 900 ms display cap followed by a logged 100 ms blank. Contact occurred within the blank at 918 ms ( 83 ms before subsequent No-Go onset). Because the software attributed the tap to the impending No-Go stimulus, the logged latency was 918 – 1,001 = -83 ms. Red dashed outlines represent the inferred on-screen duration, as stimulus offset timestamps were. **(B, D)** Extreme negative latencies beyond -100 ms (N = 11, reaching -511 ms). These require an unlogged gap: an unrecorded 435 ms delay occurred between the end of the blank and No-Go onset (purple dashed outline), while contact landed at 929 ms after the timed-out Go. Times in the listings reflect elapsed milliseconds from the preceding Go onset. All 21 events fall below the standard 100 ms post-stimulus floor and were automatically excluded from commission-error tallies without requiring post-hoc data adjustments (see Supplementary Results SR6). Abbreviations: RT, reaction time; SP-MS, secondary progressive multiple sclerosis.

Supplementary Figure S5

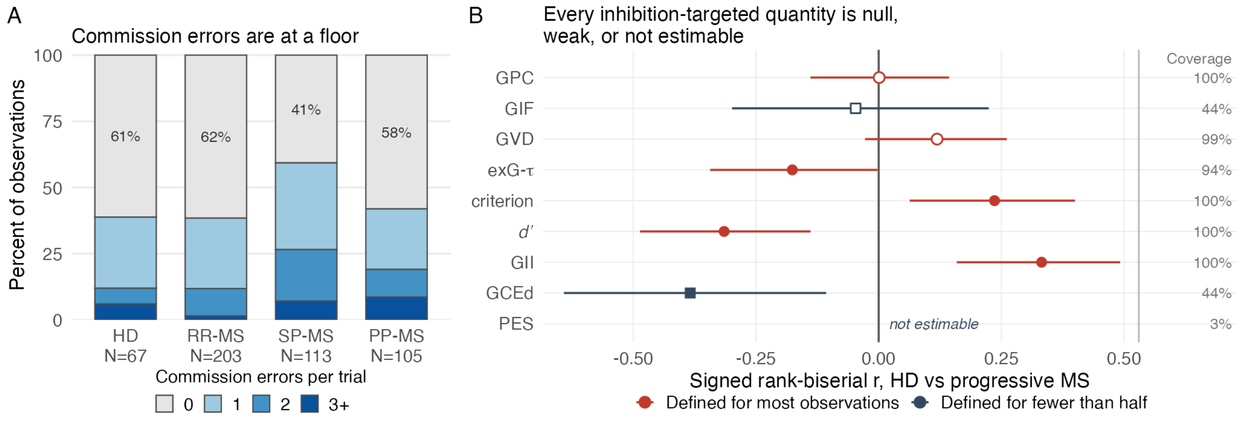


**Supplementary Figure S5. Floor effects in the equiprobable design render inhibition metrics null, weak, or unestimable. (A)** Distribution of raw commission errors per patient-hand under the primary attribution rule, stacked to 100% within each diagnostic group. Raw counts are presented instead of logarithmic rates to illustrate the pervasive floor effect: 55.9% of all 488 observations recorded zero commission errors (a structural floor obscured by $\log_{10}$ transformations). Within-group normalization accounts for fourfold sample size disparities. While the zero-error proportion is smaller in progressive MS than in HD (consistent with the direction of the GII effect), error rates do not increase monotonically across MS disease stages. **(B)** Forest plot of nine metrics targeting inhibition and sustained attention, comparing HD to progressive MS alongside metric-level data coverage. Effect sizes represent rank-biserial correlations with 95% patient-clustered bootstrap percentile intervals (open symbols denote unadjusted $p \geq 0.05$). Directionality is retained because the sign itself is informative: GIF, GPC, GVD, and exG-τ were hypothesized to increase in progressive MS, yet three were non-significant and exG-τ was significantly negative. Visual coding distinguishes high-coverage metrics (> 50% estimable) from low-coverage ones (< 50%), separating genuine null findings of preserved inhibitory control from floor-induced measurement failures. Post-error slowing (PES) is designated "not estimable" due to extreme missingness (N = 1 in HD; see Section 2.4). Abbreviations: criterion, response bias; d', sensitivity; exG-τ, ex-Gaussian τ; GCEd, Go/No-Go commission-error reaction-time difference; GIF, Go/No-Go inhibitory fatigue; GII, Go/No-Go inhibition impairment; GPC, Go/No-Go prepotency cost; GVD, Go/No-Go vigilance decrement; HD, healthy donor; N, number of first-trial patient-hand observations; PES, post-error slowing; PP-MS, primary progressive multiple sclerosis; r, rank-biserial correlation; RR-MS, relapsing-remitting multiple sclerosis; SP-MS, secondary progressive multiple sclerosis.

#### Supplementary Tables

**Supplementary Table S1.** Cohort characteristics for the four analysis groups and the three non-MS groups held out of the group comparisons. Values are median [IQR] unless stated. Age and disease duration are taken at each participant's first analyzable trial and EDSS (Expanded Disability Status Scale) is the closest clinician exam within ±7 days of it. Trial counts are trials passing quality control; first-trial observations are one trial per participant per hand, selected identically in every group.

**Supplementary Table S2.** Group separation for the ten reported outcomes. Median per diagnostic group, the Kruskal-Wallis omnibus test with its Benjamini-Hochberg-adjusted p, the HD-versus-progressive-MS rank-biserial correlation, and the severity gradient. Supports Section 3.2 and main-text Figure 3.

**Supplementary Table S3.** All 60 pairwise group contrasts for the ten reported outcomes. The six group pairs for each of the ten outcomes, as Mann-Whitney rank-biserial correlations with p-values Holm-adjusted within an outcome, together with each outcome's omnibus q and a column marking the contrasts that survive the protected post-hoc rule applied in main-text Figure 3A. Supports Section 3.2 and main-text Figure 3.

**Supplementary Table S4.** Between-session test-retest reliability of the ten reported outcomes and of the GNG-C composite. Two-way consistency, single-measures intraclass correlation coefficients (ICC), estimated as variance ratios from a mixed model with random patient-hand and trial effects, in two retest windows: the primary window (the first three trials per patient-hand within 90 days of that patient-hand's first trial) and a secondary window (up to ten trials per patient-hand, no interval restriction); the coefficients from the exploratory unrestricted window over the entire follow-up are given in Supplementary Results SR5. Both require at least two trials per patient-hand. Confidence intervals are the 2.5th and 97.5th percentiles of 2,000 bootstrap replicates that resample participants, so a participant's two hands are resampled together. Bands follow the conventional thresholds (poor < 0.50, moderate 0.50–0.74, good 0.75–0.89, excellent ≥ 0.90). Rows are ordered by tier and then by descending primary-window ICC. GNG-C is the linear predictor of the stepwise ordinal staging model, scored with fixed weights.

**Supplementary Table S5**. Between-session reliability of the GNG-C and MSC-2 composites, on each task's own retest cohort and on matched patient-hands. The estimator, window definitions and bootstrap procedure are those of Supplementary Table S4, applied to both tasks. Neither composite is refit: GNG-C is the linear predictor of the Go/No-Go stepwise ordinal model and MSC-2 that of the two-predictor motor sequencing model, each scored with its published weights. The "own retest cohort" rows use every patient-hand a task contributes to the window; the "matched patient-hands" rows are restricted to the patient-hands present in both tasks' windows, which is what makes the two coefficients comparable. Because the MSC-2 coefficients are recomputed here under this paper's mixed-model estimator rather than quoted, they replicate the published 90-

day value (0.716 against 0.70) and run lower over ten trials (0.699 against 0.77), where the two estimators diverge.

**Supplementary Table S6.** Commission-error attribution: the three-rule comparison – raw event stream, primary narrow rule, strict rule – with the proportion of observations recording no commission error, the HD-versus-progressive-MS separation and the between-session ICC under each, and the rank agreement between rules. Supports SM2, SM3 and SR2.

**Supplementary Table S7.** Full correlation matrix: every outcome and GNG-C against every clinical, functional and imaging measure tested, with N, the raw and the oriented rho, the adjusted p-value, and a column marking which cells main-text Figure 5A draws. The four regional lesion loads are the rows the figure omits as a display choice. Supports Section 3.5, main-text Figure 5 and SR7.

**Supplementary Table S8.** Nested model comparison for disease staging. Four ordinal staging models – motor sequencing alone, Go/No-Go alone, both, and a stepwise reduction of the combined model with the two MSC-2 predictors forced in – with predictor count, training and held-out accuracy, adjacent accuracy, held-out concordance and AIC. The concordance column is the single reference split; the median and 10th–90th percentile over the 500 splits are given in Section 3.6 and SR8 and drawn in main-text Figure 6B. Supports Section 3.6, main-text Figure 6 and SR8.

**Supplementary Table S9.** Incremental explained variance, with the reciprocal increment. All 40 outcome-by-biomarker increments over the motor sequencing baseline – the four outcomes of Section 2.5 crossed with the ten reported Go/No-Go outcomes – each beside its reciprocal increment and the fold difference between them. Supports Section 3.6, main-text Figure 6 and SR8.

**Supplementary Table S10.** Sensitivity of the staging comparison to the split protocol. The comparison of Supplementary Table S8 repeated under the two split-protocol choices, the stratification variables and the handling of the healthy participants, as medians over 200 reseeds per protocol with the increment from adding Go/No-Go and the proportion of splits in which that increment is negative. Supports Section 3.6, main-text Figure 6 and SR8.

**Supplementary Table S11.** Stimulus composition and sequential structure: the aggregate Go proportion with its binomial test, the under-dispersion analysis establishing that balance is enforced by design, the generating mechanism, order within the balanced set, the lag-1 transition matrix against both the Bernoulli and the without-replacement null, the continuation past the first set, and trial length. Supports SM1 and SR3.

**Supplementary Table S12.** Go run-length distribution: runs observed and percentage of runs at each length from 1 to 12. Supports SR3.

**Supplementary Table S13.** Outcome screen: all 16 candidates with tier, coverage, HD-versus-progressive-MS separation, severity gradient, test-retest reliability, the largest correlation with an outcome already in the reported family, and the tier assignment with its reason. Main-text Table 1 gives the definitions; this table is the evidence behind each tier. Supports SM3 and SR6.

**Supplementary Table S14.** GNG-C across all seven diagnostic groups. N, participants, median age and median GNG-C per group; ages are medians over observations, the unit GNG-C and the omnibus test are computed on, so a participant contributing both hands counts twice and the medians differ by a few tenths of a year from the per-participant ages of Supplementary Table S1. Supports SM7 and SR1.

**Supplementary Table S15.** Pairwise GNG-C contrasts among the seven diagnostic groups. All 21 pairwise contrasts with rank-biserial effect size, unadjusted p and Benjamini-Hochberg q. Supports SM7 and SR1.

#### Abbreviations

| Abbreviation | Expansion |
|---|---|
| 50/50GNG | equiprobable Go/No-Go; the continuous vigilance and choice-reaction task |
| 9HPT | 9-hole peg test |
| AIC | Akaike information criterion |
| CI | confidence interval |
| CIS/RIS | clinically or radiologically isolated syndrome |
| CombiWISE | Combinatorial Weight-Adjusted Disability Scale |
| criterion | response bias (signal detection) |
| d' | sensitivity (signal detection) |
| EDSS | Expanded Disability Status Scale |
| exG-μ | ex-Gaussian μ |
| exG-σ | ex-Gaussian σ |
| exG-τ | ex-Gaussian τ |
| GCEd | Go/No-Go commission-error reaction-time difference |
| GIF | Go/No-Go inhibitory fatigue |
| GII | Go/No-Go inhibition impairment |
| GME | Go/No-Go method error rate |
| GNG-C | Go/No-Go composite; the five-predictor ordinal composite of this study |
| GOI | Go/No-Go omission impairment |
| GPC | Go/No-Go prepotency cost |
| GRT | Go reaction time |
| GRTa | Go/No-Go adjusted reaction time |
| GVD | Go/No-Go vigilance decrement |
| HD | healthy donor |
| ICC | intraclass correlation coefficient |
| IQR | interquartile range |
| iSMPT | individualized sensory-motor processing threshold |
| KS | Kolmogorov–Smirnov |
| MS | multiple sclerosis |
| MSC-2 | two-predictor motor sequencing composite of the MST |
| MST | Motor Sequencing Test |

| Abbreviation | Expansion |
|---|---|
| NeurEx | digitalized neurological examination; NeurEx total is its total score |
| NeuFun-TS | Neurological Functions Test Suite |
| NIND | non-inflammatory neurological disease |
| OIND | other inflammatory neurological disease |
| OLS | ordinary least squares |
| PES | post-error slowing |
| P-MS | progressive multiple sclerosis (PP-MS + SP-MS) |
| PP-MS | primary progressive multiple sclerosis |
| QC | quality control |
| RR-MS | relapsing-remitting multiple sclerosis |
| rSDMT | randomized Symbol Digit Modalities Test |
| RT | reaction time |
| SD | standard deviation |
| SP-MS | secondary progressive multiple sclerosis |

Statistical symbols used in this supplement:

| Symbol | Meaning |
|---|---|
| C | concordance index |
| H | Kruskal-Wallis test statistic |
| q | Benjamini-Hochberg-adjusted p-value |
| r | rank-biserial correlation |
| $r_{SB}$ | Spearman-Brown split-half reliability coefficient |
| ρ | Spearman's rank correlation coefficient |
| $\Delta R^2$ | increment in explained variance |
